# Safety and immunogenicity of a SARS-CoV-2 recombinant protein vaccine with AS03 adjuvant in healthy adults: interim findings from a phase 2, randomised, dose-finding, multi-centre study

**DOI:** 10.1101/2021.10.08.21264302

**Authors:** Saranya Sridhar, Arnel Joaquin, Matthew I Bonaparte, Agustin Bueso, Anne-Laure Chabanon, Aiying Chen, Roman M. Chicz, David Diemert, Brandon J Essink, Bo Fu, Nicole A Grunenberg, Helene Janosczyk, Michael C Keefer, Doris M Rivera M, Ya Meng, Nelson L. Michael, Sonal S Munsiff, Onyema Ogbuagu, Vanessa N Raabe, Randall Severance, Enrique Rivas, Natalya Romanyak, Nadine G Rouphael, Lode Schuerman, Lawrence D Sher, Stephen R Walsh, Judith White, Dalia von Barbier, Guy de Bruyn, Richard Canter, Marie-Helene Grillet, Maryam Keshtkar-Jahromi, Marguerite Koutsoukos, Denise Lopez, Roger Masotti, Sandra Mendoza, Catherine Moreau, Maria Angeles Ceregido, Shelly Ramirez, Ansoyta Said, Fernanda Tavares-Da-Silva, Jiayuan Shi, Tina Tong, John Treanor, Carlos A Diazgranados, Stephen Savarino

## Abstract

**Background:** This study evaluated the safety and immunogenicity of an AS03-adjuvanted SARS-CoV-2 recombinant protein candidate vaccine, CoV2 preS dTM.

**Methods:** This Phase 2, modified double-blind, parallel-group study (NCT04762680) was conducted in adults, including those at increased risk of severe COVID-19. Participants were randomised 1:1:1, stratified by age (18–59/≥60 years), rapid serodiagnostic test (positive/negative) and high-risk medical conditions (yes/no), to receive two injections (day [D]1 and D22) of 5μg, 10μg or 15μg of CoV2 preS dTM antigen with fixed AS03 content. Interim safety and reactogenicity results (to D43) and neutralising antibodies (NAbs) against the D614G variant are presented (primary objectives).

**Findings:** Of 722 participants enrolled and randomised between 24 February and 8 March 2021, 721 received ≥1 injections (5μg, n=240; 10μg, n=239; 15μg, n=242). Four participants reported unsolicited immediate adverse events (AEs), two were vaccine-related (investigator assessment). Five participants reported seven vaccine-related medically-attended AEs. No vaccine-related serious AEs and no AEs of special interest were reported. Solicited reactions (local and systemic) were reported at similar frequencies between study groups; these were mostly mild to moderate and transient, with higher frequency and intensity post-injection 2 than post-injection 1. In SARS-CoV-2 naïve participants at D36, 96·9%, 97.0% and 97·6% of participants had ≥4-fold-rise in NAb titres from baseline in the 5μg-, 10μg- and 15μg-dose groups, respectively. NAb titres increased with antigen dose in younger (GMTs: 2954, 3951 and 5142 for 5μg-, 10μg- and 15μg-dose groups) but not older adults (GMTs: 1628, 1393 and 1736, respectively). NAb titres in non-naïve adults after one injection were higher than titres after two injections in naïve adults.

**Interpretation:** Two injections of CoV2 preS dTM-AS03 demonstrated acceptable safety and reactogenicity, and robust immunogenicity in SARS-CoV-2 naïve and non-naïve adults. These results informed antigen dose selection for progression to Phase 3 evaluation of primary and booster vaccination.

## Introduction

Coronavirus disease 2019 (COVID-19), caused by severe acute respiratory syndrome coronavirus 2 (SARS-CoV-2), has inflicted unprecedented morbidity and mortality worldwide and continues to devastate global health and economies more than a year since its emergence.^1-3^

Extraordinary efforts in the development, manufacturing and distribution of COVID-19 vaccines have led to several vaccines with emergency use designation,^4^ and more recently full approval.^5^ Despite this remarkable progress, continued efforts to develop vaccines are necessary to meet global demand by offering alternative vaccine choices for optimal benefit-risk across diverse populations and providing broader protection against emerging variants often in the face of varying patient and provider preference.^6^

Sanofi Pasteur, in collaboration with GlaxoSmithKline, have developed an adjuvanted SARS-CoV-2 recombinant protein vaccine (CoV2 preS dTM) using a baculovirus expression vector system (BEVS) to express stabilised SARS-CoV-2 pre-fusion Spike (S) antigen (preS dTM).^7^ The use of an adjuvanted vaccine formulation offers advantages of dose-sparing and greater breadth of protection.^8^ In a Phase 1/2 study, the safety and immunogenicity of the CoV2 preS dTM candidate vaccine adjuvanted with AS03 (GlaxoSmithKline) and AF03 (Sanofi Pasteur) at two antigen dose levels (effective antigen doses administered, 1·3 and 2·6 μg) was evaluated in healthy adults aged 18 years or older.^7^ Interim data from that study enabled selection of a two-injection schedule and the AS03 adjuvant for further clinical development. However, lower than expected antibody responses, particularly in older adults (≥60 years), and higher than expected reactogenicity after the second vaccination was observed. This was hypothesised to be due to lower than planned antigen doses and higher than anticipated host-cell protein content in the formulations tested.^7^ Optimised CoV2 preS dTM formulations with increased antigen content and lower host-cell protein content have been developed and are being evaluated in further clinical development.

The current Phase 2 study evaluated the safety, reactogenicity and immunogenicity of three optimised formulations of AS03-adjuvanted CoV2 preS dTM containing 5, 10 or 15 μg of antigen, administered as two intramuscular injections 21 days apart, to determine progression and optimal antigen dose for Phase 3 efficacy evaluation.

## Methods

### Study design and participants

This is an ongoing Phase 2, randomised, modified double-blind, parallel group, dose-ranging study (clinicaltrials.gov NCT04762680). The study is being conducted in 20 clinical research centres in the USA and Honduras, with a planned duration of approximately 13 months. Here, we present interim safety and reactogenicity data up to study day (D) 43, three weeks after the second vaccination, and immunogenicity data up to D36.

Adults, aged 18 years and older, were eligible. Women who were pregnant or lactating, or, for those of childbearing potential, not using an effective method of contraception or abstinence, were excluded. To allow evaluation of vaccine performance in high-risk groups, individuals with pre-existing medical conditions, those who were immunocompromised (except those with recent organ transplant or chemotherapy) and those with a potentially increased risk for severe COVID-19,^9^ were eligible for participation in the study. The inclusion/exclusion criteria are described in detail in the Appendix (**Appendix – Supplementary Methods 1**).

The study was undertaken in compliance with the International Conference on Harmonisation (ICH) guidelines for Good Clinical Practice and the principles of the Declaration of Helsinki. The protocol and amendments were approved by applicable Independent Ethics Committees/Institutional Review Boards, including the Advarra Institutional Review Board for the USA, and the regulatory agencies as per local regulations. Written informed consent was obtained from the participants before any study procedures were performed.

### Randomisation and masking

Participant randomisation was stratified by age group (18–59 years and ≥60 years), by positivity for antibodies to SARS-CoV-2 S antigen using a rapid serodiagnostic test (positive/negative; COVID-19 IgG/IgM Rapid Test Cassette; Healgen Scientific, Houston, TX, USA) and by high-risk medical conditions (yes/no). Participants were randomly assigned, using an interactive response technology system, in a 1:1:1 ratio to receive two intramuscular injections, 21 days apart (D1 and D22), of one of the three candidate vaccine formulations containing 5 μg (low-dose), 10 μg (medium-dose) or 15 μg (high-dose) of CoV2 preS dTM antigen with a fixed dose of AS03 adjuvant. A subset of 120 participants who tested negative by the SARS-CoV-2 rapid serodiagnostic test was additionally stratified by age and study group and randomly assigned to provide samples for assessment of cell-mediated immunity (CMI) and mucosal antibodies.

This study had a modified double-blind design, such that all participants and outcome assessors were masked to group assignments and unblinded study staff involved in vaccine preparation were not involved in safety outcome assessments.

### Procedures

The recombinant protein antigen CoV2 preS dTM was produced using a Sanofi Pasteur proprietary insect cell baculovirus expression vector system, based on S protein sequence from the Wuhan-Hu-1 reference strain, as previously described.^7^ The AS03 Adjuvant System (GlaxoSmithKline Vaccines, Rixensart, Belgium) is an oil-in-water emulsion containing 11·86 mg α-tocopherol and 10·69 mg squalene per 0·5 mL vaccine dose, described previously.^7,10^ The AS03-adjuvanted CoV2 preS dTM vaccine formulations were presented in two separate vials: a multi-dose vial containing AS03 and a single-dose vial containing one of the three antigen dose solutions. An equal volume of the adjuvant emulsion was added to the vial containing the antigen and mixed prior to injection. Vaccinations (0.5 mL per dose) were administered by qualified and trained study personnel by intramuscular injection into the deltoid region of the upper arm.

Blood samples and nasopharyngeal swabs were collected before each vaccination for determination of SARS-CoV-2 naïve/non-naïve status. SARS-CoV-2 naïve or non-naïve status at D1 and/or D22 was determined for all participants on serological samples using ELECSYS electrochemiluminescence immunoassays for anti-SARS-CoV-2 anti-S (S-ELECSYS; Elecsys Anti-SARS-CoV-2 S, Roche, Indianapolis, IN, USA EUA202698) on D1 only and anti-nucleocapsid (N) (N-ELECSYS; Elecsys Anti-SARS-CoV-2, Roche, EUA200514) on D1 and D22, and on nasopharyngeal swabs using nucleic acid amplification tests (NAAT; Abbott RealTime SARS-CoV-2 assay, EUA200023) on D1 and D22, following the manufacturer’s instructions. SARS-CoV-2 naïve participants at D1 and D22 (naïve D1+D22) were those who tested negative for S-ELECSYS at D1 and negative for both N-ELECSYS and NAAT at both timepoints; the non-naïve subset of participants tested positive for at least one of the three tests at D1 and/or D22 (non-naïve D1/D22) (**Appendix – Methods 2**).

Blood samples were collected from participants at all study visits for immunogenicity assessments. SARS-CoV-2 neutralising antibody (NAb) titres against the D614G variant and the Beta (B.1.351) variant were measured using a pseudovirus neutralisation (PsVN) assay performed at Monogram Biosciences LabCorp (South San Francisco, CA, USA). The assay, based on previously described methodologies, measured NAb responses using HIV-1 pseudovirions expressing the full-length S protein of the parental D614G variant or the Beta variant.^11-13^ Neutralisation by SARS-CoV-2 antibodies was measured by inhibition of luciferase activity in the target cells. Titres were calculated as the reciprocal of the serum dilution (dil) resulting in 50% neutralisation. Binding antibody (BAb) profiles were assessed by measuring SARS-CoV-2 anti-S protein IgG antibodies with an indirect enzyme-linked immunosorbent assay (ELISA) (Nexelis, Laval, Quebec, Canada), as described previously.^7^ The reference standard was human serum with known concentration of anti-S protein IgG antibodies; quantitative results were reported in EU/mL.

Whole blood samples were stimulated *ex vivo* with SARS-CoV-2 S Antigen (Spike-GCN4, Nexelis), using the TruCulture system (Rules-Based Medicine [RBM], Austin, TX, USA) as described previously.^7^ A microsphere-based multiplex immunoassay (TruCulture OptiMAP™ assay, RBM) was used to evaluate specific concentrations of interferon-γ [IFN γ], tumour necrosis factor-α [TNF-α], interleukin [IL]-2, IL-4, IL-5 and IL-13 on validated cytokine profiling panels. Reaction plates were analysed on a Luminex® platform and cytokine concentrations were calculated by adapted software (RBM) using a standard curve for specific cytokine production at each time point.

Participants were provided a diary card to capture solicited and unsolicited adverse events (AEs) for up to 21 days after the second injection; serious adverse events (SAEs), adverse events of special interest (AESIs) and medically attended adverse events (MAAEs) were collected over the duration of the study.

### Outcomes

The primary safety objective was to describe the safety profile in all participants, for each candidate vaccine formulation. In this interim analysis, we describe primary safety endpoints up to D43 (21 days post-injection 2). Safety endpoints included the occurrence of immediate unsolicited systemic AEs occurring within 30 minutes of each injection; solicited injection site reactions (injection site pain, erythema and swelling) and solicited systemic reactions (fever, headache, malaise, myalgia, arthralgia and chills) up to seven days after each injection; unsolicited AEs reported up to 21 days after each injection; and MAAEs, serious adverse events (SAEs) and AESIs documented throughout the study. AESIs included anaphylactic reactions, generalized convulsions, thrombocytopenia and potential immune-mediated disorders.^14^

The primary immunogenicity objective was to describe the NAb response to the D614G variant in SARS-CoV-2-naïve participants (naïve D1+D22 subset) 14 days after the second vaccination (D36) for each study group. NAb and BAb responses were measured in all participants on D1, D22, and D36 to D614G. NAb responses to the Beta variant were measured at D36. Antibody responses were described based on geometric mean titres (GMTs) or geometric mean concentration (GMCs) for NAb and BAb antibodies respectively, the percentage of participants with ≥2-fold or ≥4-fold rise in antibody titres at each post-vaccination time point and the percentage of responders. Pre-vaccination titres below the assay lower limit of quantification (LLOQ; 1:40) were assigned a value of half of the LLOQ. Among participants who had NAb titres below the LLOQ at baseline, responders were defined as those with ≥2-fold increase in titres at a pre-defined post-vaccination time point. In participants with baseline titres above LLOQ, responders were those with a ≥4-fold increase in titres post-vaccination relative to D1. The geometric means of individual titre ratios (GMTRs), post-vaccination to pre-vaccination (D1), were described for each treatment group, following each injection. For cell-mediated immune responses, fold-rises for individual cytokines at D36/D1 and D22/D1 were calculated by dividing the post-vaccination (D22 or D36) by the pre-vaccination (D1) measurement; the ratios of fold-rises for cytokine pairs (e.g. IFN-γ to IL-4) and their 95% confidence intervals (CIs) were then computed.

NAb and BAb were also measured in a panel of human convalescent serum samples (Sanguine Biobank, iSpecimen, and PPD; n=79 samples) using the same assays as used on the participant serum samples, in the same laboratory and within the same timeframe. Convalescent samples were obtained from donors who had recovered from COVID-19 (with clinical severity ranging from mild to severe) and were asymptomatic at the time of sample collection, as described previously.^7^ Ratios of vaccine-induced antibody titres to convalescent serum titres were calculated for each antigen-dose group, by age group.

### Statistics

All planned analyses were descriptive. It was estimated that a sample size of 130 evaluable SARS-CoV-2 naïve participants per group would enable a minimum observed GMT ratio between vaccine groups of 0.73, assuming a true GMT ratio of 1 and a standard deviation of 0.67 (estimated for the PsVN assay) with 95% probability. Assuming an attrition rate of 15% and capping the proportion of those testing positive by the SARS-CoV-2 rapid serodiagnostic test at 20% of the study population, a total study size of 720 participants (240 in each group) was planned.

The full analysis set (FAS) comprised all participants who received at least one study injection; those in the FAS were analysed according to the vaccine group to which they were randomised. The safety analysis set (SafAS) included all randomised participants who received at least one dose of the study vaccine and were analysed according to the vaccine actually received. Immunogenicity was assessed in participants from the per-protocol analysis set (PPAS), a subset of participants in the FAS who received both injections, provided blood samples at D1 and D36, did not have pre-specified protocol deviations for PPAS and did not receive an authorized COVID-19 vaccine prior to D36.

Primary immunogenicity endpoints were assessed in the SARS-CoV-2 naïve participant subset of the PPAS (PPAS naïve D1+D22); secondary immunogenicity endpoints were assessed in SARS-CoV-2 naïve and non-naïve participant subsets of the PPAS and FAS. CMI was analysed in a randomly assigned subset of the PPAS (CMIAS).

The 95% CIs for the GMTs/GMCs and GMTRs were calculated using normal approximation of log-transformed titres. The 95% CIs for the percentage of participants or responders were based on the Clopper-Pearson method.^15^ The 95% CIs for the differences in the percentage of participants with ≥2-fold or ≥4-fold rise and the percentages of responders were calculated using the Newcombe-Wilson score method without continuity correction.^15^ Statistical analyses were performed using SAS® Version 9.4 or later.

### Results

A total of 722 participants were enrolled and randomized in the study between 24 February 2021 and 8 March 2021; 721 received at least one injection (low-dose, n=240; medium-dose, n=239; high-dose, n=242). A total of 22 randomised participants discontinued the study by D43, none due to an AE (**Appendix – Figure S1**). Participant baseline demographic characteristics in the SafAS were balanced overall across treatment groups, and across age strata. Participants were 18 to 95 years old (18–59 years, n=360; ≥60 years, n=361) (**Table 1**); 33 were aged ≥75 years. Over 60% of participants had ≥1 high-risk medical condition (FAS; **Appendix – Table S1**).

**Table 1:** Participant demographic characteristics (SafAS)

|  | Low-dose (5 µg)<br>N=240 | Medium-dose (10 µg)<br>N=240 | High-dose (15 µg)<br>N=241 |
| --- | --- | --- | --- |
| Sex |  |  |  |
| Male | 117 (48.8%) | 126 (52.5%) | 119 (49.4%) |
| Female | 123 (51.3%) | 114 (47.5%) | 122 (50.6%) |
| Age, years |  |  |  |
| Mean (SD) | 53.8 (15.3) | 53.5 (14.8) | 53.1 (15.9) |
| Range | 20.0–92.0 | 18.0–88.0 | 19.0–95.0 |
| BMI, kg/m<sup>2</sup> |  |  |  |
| Mean (SD) | 28.4 (5.6) | 28.8 (5.9) | 28.7 (6.0) |
| Country |  |  |  |
| United States | 192 (80.0%) | 193 (80.4%) | 196 (81.3%) |
| Honduras | 48 (20.0%) | 47 (19.6%) | 45 (18.7%) |
| Race |  |  |  |
| White | 156 (65.0%) | 150 (62.5%) | 155 (64.3%) |
| American Indian or Alaska Native | 22 (9.2%) | 24 (10.0%) | 20 (8.3%) |
| Black or African American | 13 (5.4%) | 23 (9.6%) | 20 (8.3%) |
| Asian | 13 (5.4%) | 10 (4.2%) | 10 (4.1%) |
| Native Hawaiian or other Pacific Islander | 2 (0.8%) | 1 (0.4%) | 2 (0.8%) |
| Multiple | 5 (2.1%) | 2 (0.8%) | 4 (1.7%) |
| Not reported or unknown | 29 (12.1%) | 30 (12.5%) | 30 (12.4%) |
| Ethnicity |  |  |  |
| Hispanic or Latino | 68 (28.3%) | 68 (28.3%) | 67 (27.8%) |
| Not Hispanic or Latino | 170 (70.8%) | 172 (71.7%) | 173 (71.8%) |
| Not reported or unknown | 2 (0.8%) | 0 | 1 (0.4%) |
| Baseline SARS-CoV-2 rapid serodiagnostic test |  |  |  |
| Negative | 219 (91.3%) | 218 (90.8%) | 219 (91.0%) |
| Positive | 21 (8.8%) | 22 (9.2%) | 22 (9.1%) |

Data are n (%) or mean (SD)
SafAS = safety analysis set.

The proportion of participants reporting at least one solicited adverse reaction (injection site or systemic) in the first seven days after any vaccination was similar between treatment groups (low- and high-dose groups, each 91·2%; medium-dose group, 89·9%), tended to be higher in younger adults (range across groups, 92·4–95·7%) than in older adults (86·9–89·1%), and after the second vaccination (82·3–86·3%) compared to the first vaccination (76·4–80·7%) (**Figures 1A and 1B; Appendix – Table S2**). The proportion of participants reporting at least one solicited reaction of grade 3 intensity after any injection was similar between treatment groups (range across groups [all age strata], 18·8–21·8%). The most frequently reported injection site reaction (all age strata) was injection site pain (range across groups, any intensity:81·9–83·7%; Grade 3: 5·0–6·3%). The most frequently reported solicited systemic reactions (any intensity, all age strata) were: malaise (range across groups, 58·0–61·1%), headache (56·1–57·1%) and myalgia (50·6–60·1%) and, for grade 3 intensity, malaise (8·4–13·0%) (**Appendix – Table S2**; for data by age group see **Figure 1B**). Grade 3 solicited injection site and systemic reactions were transient, with most occurring on the day of or the day after vaccination (*Appendix – Figure S2*) and resolving within two days without requiring medical attention.

**Figure 1:**
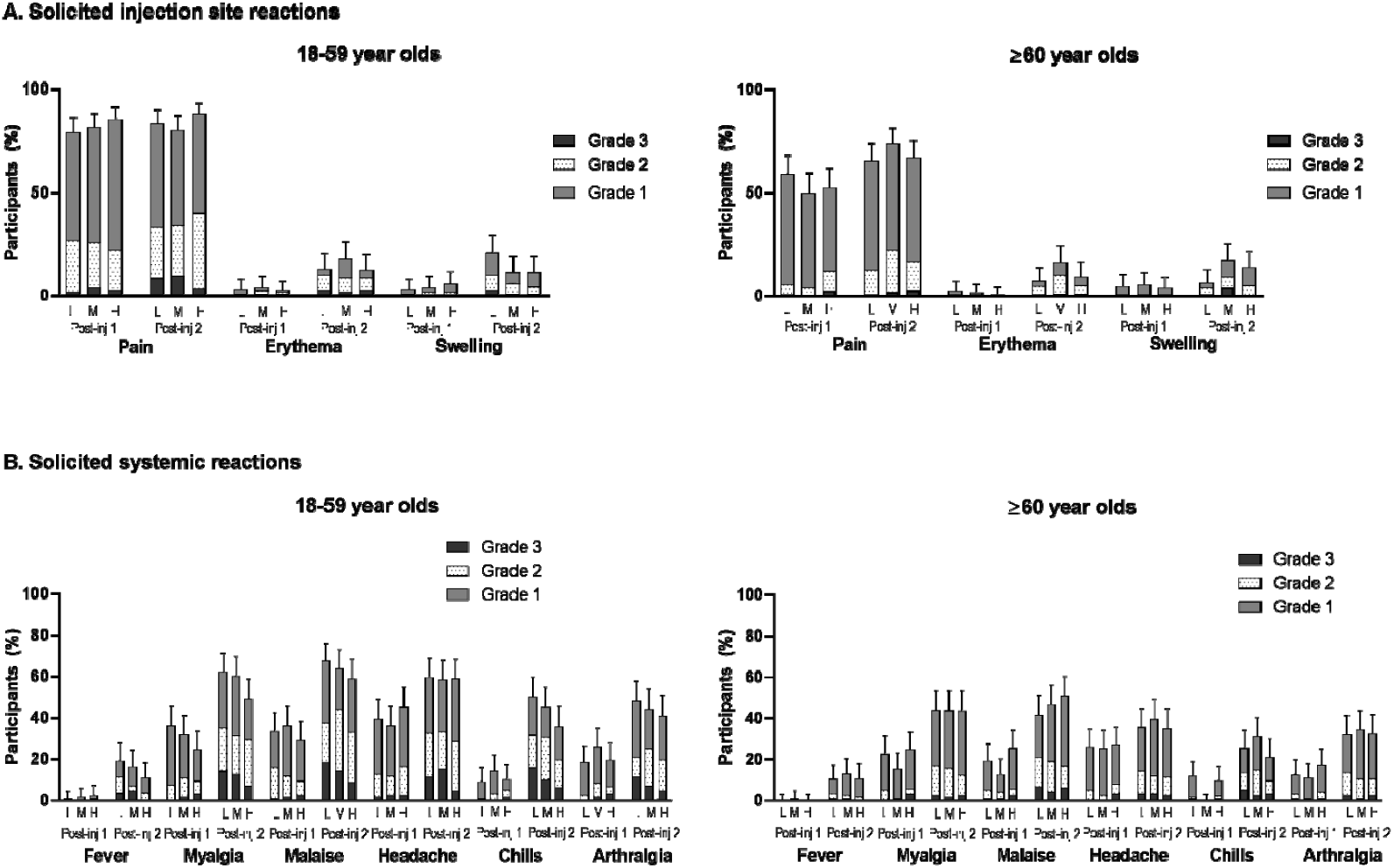
Solicited injection site (A) and systemic adverse reactions (B) after each injection for low-, medium- and high-antigen dose, by age group (SafAS) L = low-dose group (5 μg). M = medium-dose group (10 μg). H = high-dose group (15 μg). Post-inj = post-injection.

Overall, there were four immediate unsolicited systemic non-serious AEs reported in four participants (**Appendix – Table S2**): two assessed by the investigator to be vaccine-related (grade 1 lymphadenopathy [low-dose group] and grade 1 paraesthesia [medium-dose group] on the same side as the injection site) and two unrelated, grade 1 presyncope (low-dose group) and grade 3 hypertension (high-dose group). The case of lymphadenopathy resolved with medication within five days, while the others resolved spontaneously within one day. After any vaccination, the proportion of participants reporting at least one unsolicited AE was comparable across treatment groups (ranging between 32·9% and 36·9%; **Appendix – Table S2**) and following the first (range across groups, 16·3–22·0%) and second injections (range across groups, 22·8–23·6%); similar patterns were observed for unsolicited adverse reactions (ARs). The proportion of participants reporting unsolicited AEs after any injection was slightly higher among the younger than the older adults (range across groups, 36·7–41·3% versus 29·2–36·9%), while the proportion of participants reporting unsolicited ARs was similar between age strata (range across groups, 14·3–24·8% among younger adults and 13.3–14.8% among older adults). The proportion of participants with grade 3 unsolicited AEs after any injection tended to be higher in the high antigen dose group (7·9%) than in the low- (2·1%) or medium-dose (2·5%) groups; this increase with higher antigen dose was also observed with Grade 3 unsolicited ARs. The majority of unsolicited ARs were compatible with reactogenicity symptoms (**Appendix -Table S3**), were of grade 1 or grade 2 intensity, occurred within the first four days after injections and generally resolved within seven days. Six participants in the high-dose group and one participant in the low-dose group were reported to have unsolicited AEs with the MedDRA preferred terms (PTs) of elevated blood pressure, elevated systolic blood pressure, essential hypertension or hypertension. These occurred shortly after vaccination, self-resolved within 1–2 days and occurred without any other associated symptoms in all but one case; one participant with grade 3 hypertension had macular rash and headache, all assessed as related to the study vaccine, and anxiety (assessed as unrelated). COVID-19 illness was reported for four participants with the onset of illness prior to D36 (14 days after the second vaccination); one in each of the low- and medium-dose groups and two in the high-dose group. MAAEs were reported for 62/721 (8·6%) participants overall, with no clear pattern between treatment groups (**Appendix – Table S2**). Of these, 14 were grade 3 intensity, seven (reported by five participants) were assessed by the investigator as related to the vaccine (low-dose, n=2; medium-dose, n=1; high-dose, n=4) and three (not related) were assessed as serious (medium-dose, n=2; high-dose, n=1). Grade 3 MAAEs tended to be more frequent in the high-dose group (3·3%; 8/241) compared to the low- and medium-dose groups (1.7% [4/240] and 0.8% [2/240], respectively); these included two events of grade 3 hypertension assessed as related to the study vaccine, both with a duration of 2 days. No AEs led to study discontinuation, and no AESIs were reported. Four SAEs were reported (two in each of the medium-dose and high-dose groups), none of which were considered by the investigator or the sponsor to be related to the study vaccine. The safety and reactogenicity profiles were similar between participants who were SARS-CoV-2 naïve at baseline (D1) and those who were non-naïve at D1 (**Appendix - Table S2**).

Among 611 participants in the PPAS, 598 had sufficient information to determine per-protocol SARS-CoV-2 naïve status at D1 and D22: naïve, n=521 (85·2%); non-naïve, n=77 (12·6%), with similar proportions across treatment groups (naïve: n=168, n=177, n=176; non-naïve: n=28, n=26, n=23 for low-, medium- and high-dose groups, respectively).

Among SARS-CoV-2 naïve participants in the PPAS, NAb titres and BAb concentrations to the D614G variant 21 days after the first injection (D22) showed minimal increase from baseline, regardless of antigen dose group, for both age strata (**Figure 2A and Figure 3A**).After the second injection (D36), Nab titres tended to increase with antigen-dose level in the younger age group but not in the older adults; titres were higher for younger adults than for older adults within each dose group (**Figure 2A**). On D36, the proportion of responders (≥2-fold-rise in NAb titres from D1 to D36) ranged from 97·4% to 100% across treatment groups for younger adults (18–59 years) and 95·1% to 100% for older adults (≥60 years); the proportion of those with a ≥4-fold-rise in Nab titres ranged from 97·4% to 100% among younger adults and 93·9% to 96·7% among older adults (**Table 2**). Similar observations were made for D36 BAb responses (**Appendix – Table S4**). For both age strata combined, the magnitude of the NAb response after two injections (D36) was comparable across treatment groups (D36 GMTs [1/dil] ranging from 2189 [low-dose] to 2895 [high-dose group]), and comparable to titres observed in the convalescent sera panel (2140 [95% CI 1543, 2967]) (**Table 2**). The ratio of vaccine-induced NAb titres to titres in the convalescent panel was 1·38, 1·85, and 2·40 among younger adults, and 0·76, 0·65 and 0·81 among older adults for the low-, medium- and high-antigen dose groups, respectively.

**Figure 2:**
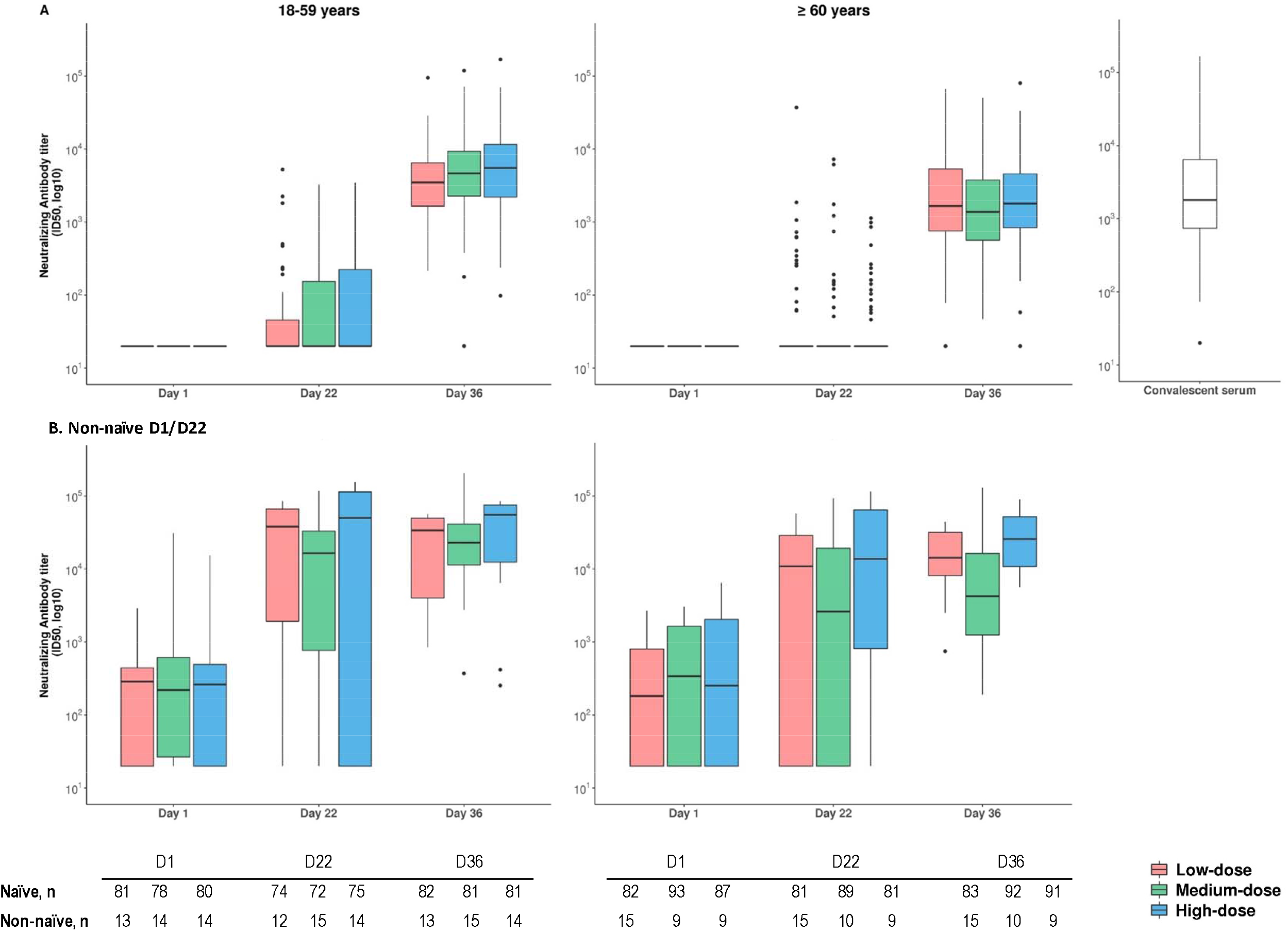
Neutralising antibody response to D614G, after each injection of low-, medium- and high-dose CoV2 preS dTM-AS03 formulations, by SARS-CoV-2 naïve status (PPAS). Footnote: Boxes indicate median and quartile ranges. Outliers are plotted as individuals points. D, study day; n, number of participants available for each endpoint are shown in the table. For the panel of convalescent sera, n=79

**Figure 3:**
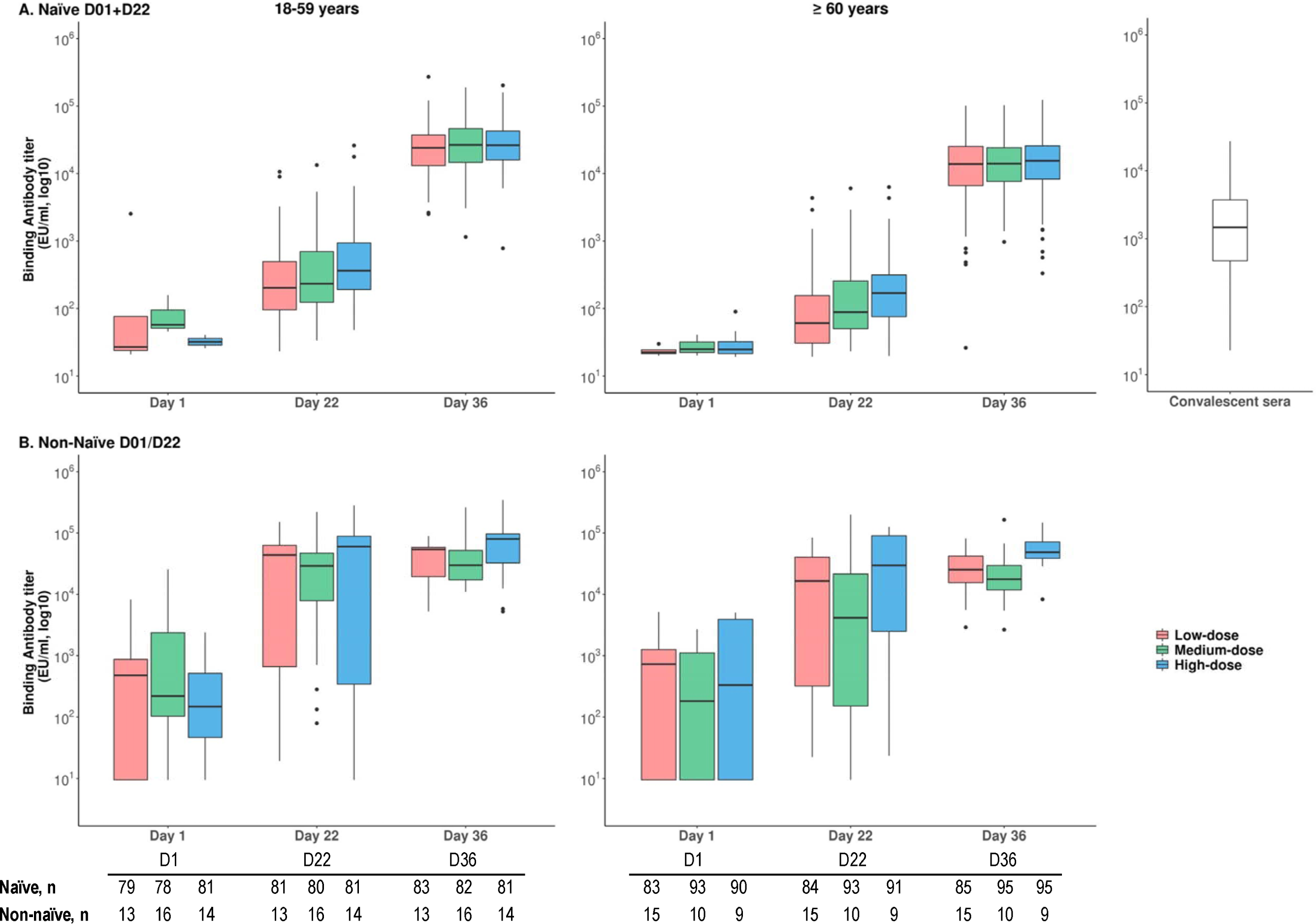
Binding antibody response to D614G, following each injection of low-, medium- and high-dose CoV2 preS dTM-AS03 formul ations, by SARS-CoV-2 naïve status. Footnote: Boxes indicate median and quartile ranges. Outliers are plotted as individuals points. D, study day; n, number of participants available for each endpoint are shown in the table. For the panel of convalescent sera, n=78

**Table 2:**
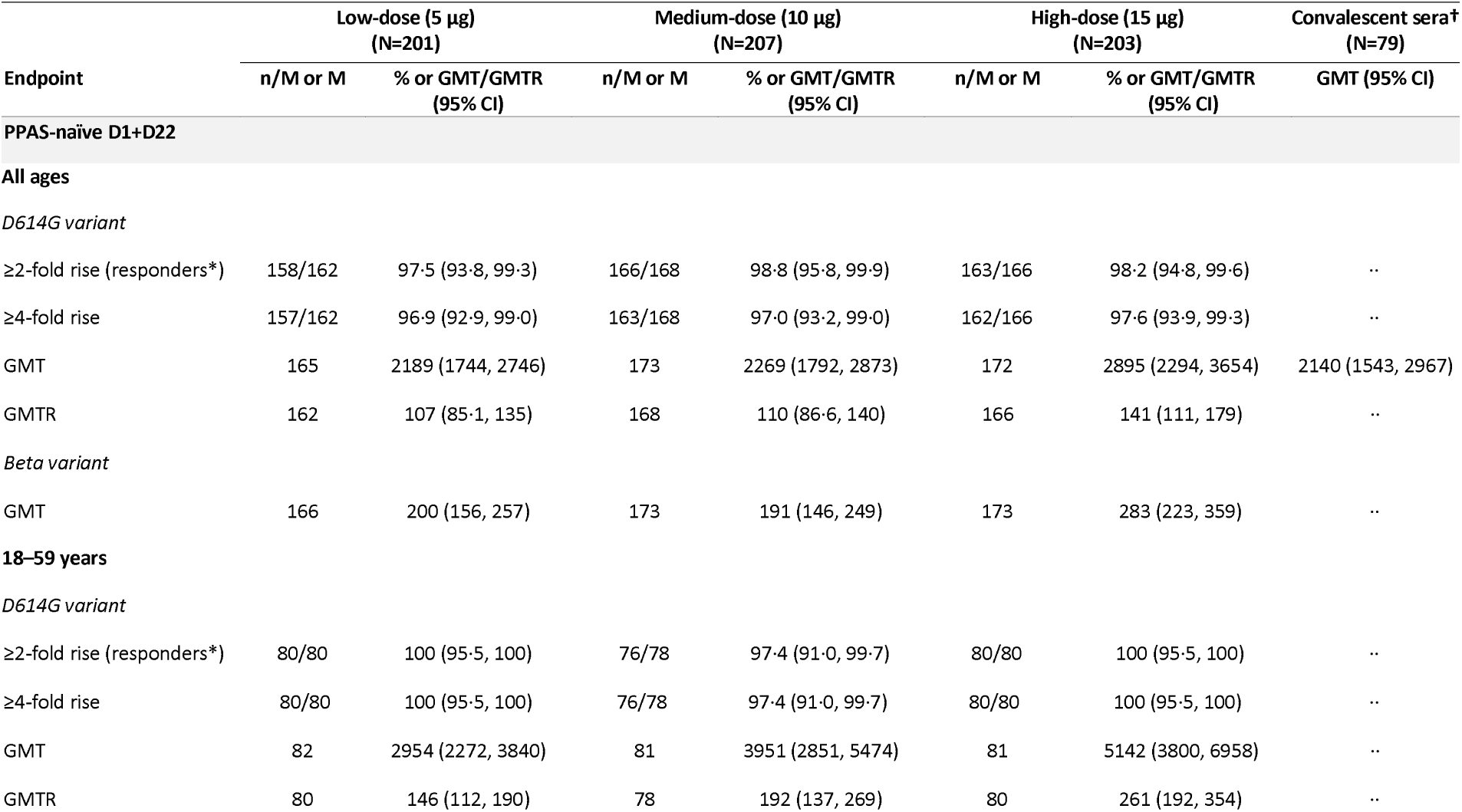
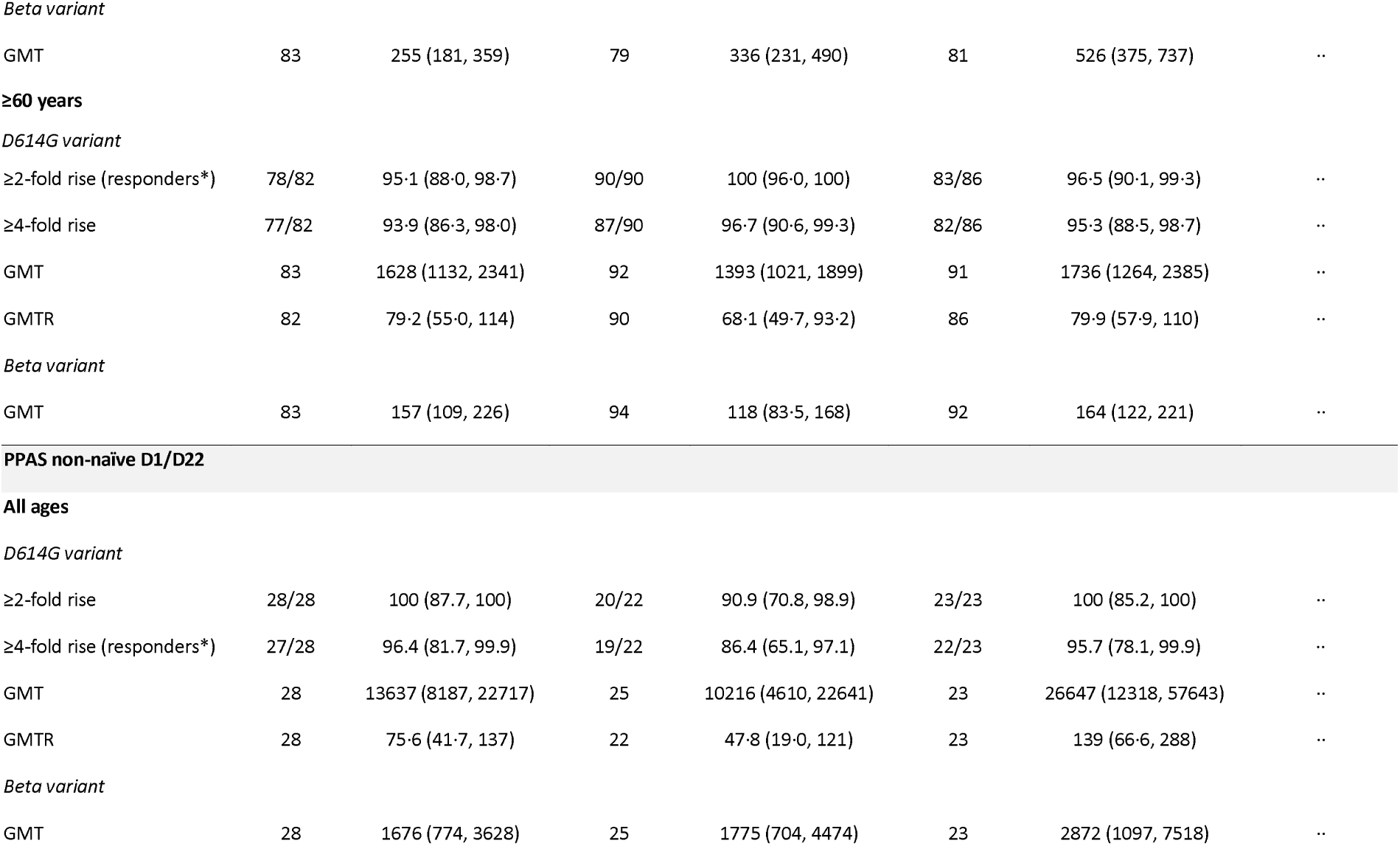
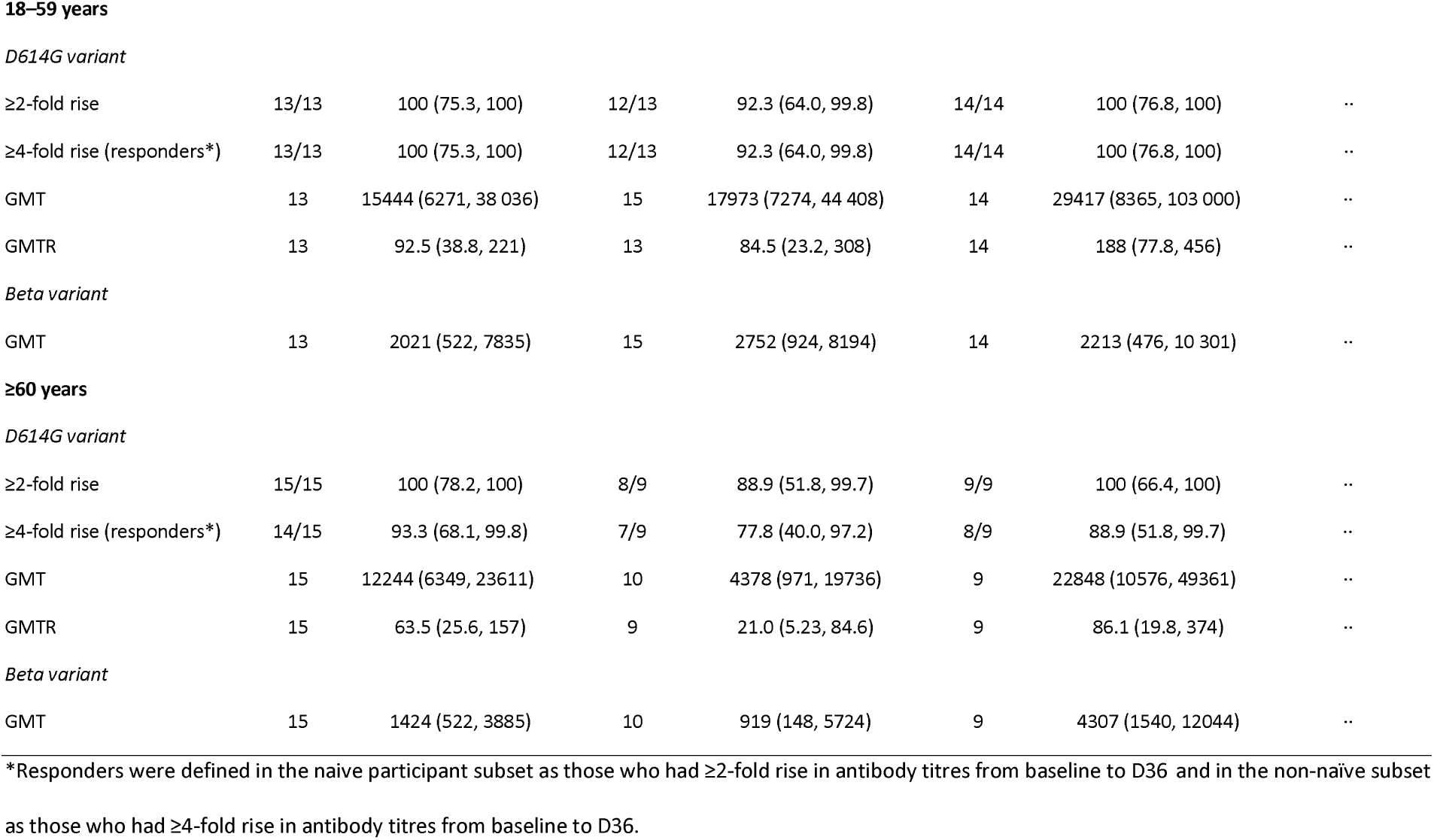
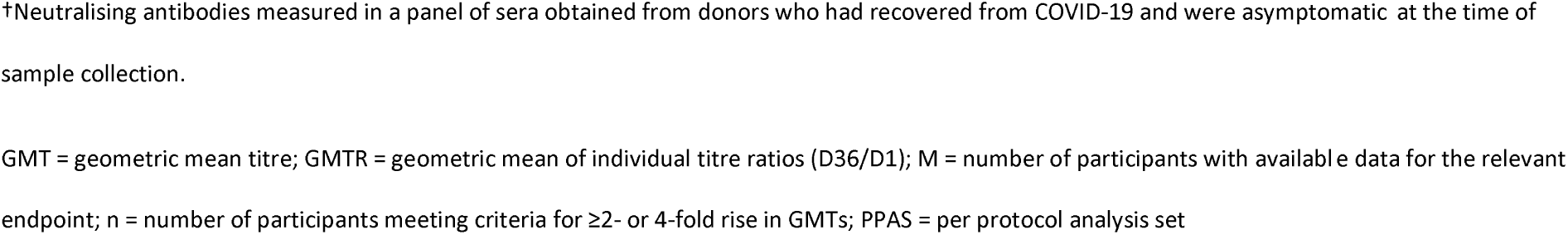
Neutralising antibody responses to D614G, and to the Beta variant at D36, by age group and by SARS-CoV-2 naïve status (PPAS)

Among SAR-CoV-2 naïve participants with at least one high risk medical condition, antibody titres to D614G variant were comparable across all antigen dose groups in both age strata. Among naïve participants without high-risk medical conditions, higher NAb titres with increasing antigen dose level was observed in younger adults (**Appendix – Table S5**). After two doses, NAb titres among those without high-risk conditions were higher than those with ≥1 high-risk medical condition in the medium- and high-dose groups, particularly in the younger age stratum, although this was not observed in the low-dose group.

In participants in the PPAS who were non-naïve on D1 and/or D22, NAb titres (**Figure 2B**) and BAb concentrations (**Figure 3B**) to D614G variant increased by more than 10-fold in both age strata 21 days after a single injection (D22) in all antigen-dose groups. Higher titres and greater fold-increases were observed in the younger age strata compared to the older age group. After the first injection, a ≥2-fold rise in NAb titres was observed for 74·1% (20/27), 59·1% (13/22), and 73·9% (17/23) participants (all ages), in the low-, medium- and high-dose groups, respectively, and a ≥4-fold rise for 66·7% (18/27), 54·5% (12/22) and 69·6% (16/23) participants, respectively. GMTs increased further after the second injection, albeit to a lesser extent, with nearly all participants in each group achieving ≥4-fold rise in NAb and BAb levels at D36 (**Table 2; Appendix – Table S5**). The D22 titres in non-naïve participants were higher across all antigen-dose groups than those achieved among naïve participants after two doses (D36) (**Figures 2 and 3**).

The NAb response to the Beta variant was assessed at D36 only. In the PPAS naïve D1+D22 participants, GMTs were comparable between the low- and medium-dose groups, and slightly higher for the high-dose group, with titres approximately 10-fold lower than for the D614G variant (**Table 2; Appendix – Figure S3**). The pattern of NAb responses to the Beta variant with age was similar to that observed with responses to the D614G variant, with higher titres in younger compared to older adults. In the non-naïve participants, the Beta variant GMTs were also comparable between the low- and medium-dose groups with higher titres for the high-dose group (**Table 2**).

Cell-mediated immunity was assessed in a subset of 104 participants: 36 participants in the low-dose group, 31 participants in the medium-dose group, and 37 in the high-dose group. Increases of IFN-γ, IL-2, and TNF-α cytokines from pre-vaccination to D22 and D36 were more robust than were the increases for IL-4, IL-5, and IL-13, suggesting no Th2 cell bias in the cell-mediated responses (**Appendix – Figure S4**).

## Discussion

In this study, two injections of the AS03-adjuvanted SARS-CoV-2 recombinant protein vaccine, CoV2 preS dTM-AS03, demonstrated an acceptable safety and reactogenicity profile, and favourable neutralising antibody and cellular immune responses in SARS-CoV-2 naïve and non-naïve adults, for all three antigen dose groups, and in both younger (18–59 years) and older (≥60 years) age strata.

No safety concerns were identified during the interim study period. In the current study, local and systemic solicited reactions (overall and grade 3 intensity) were reported more frequently after the second injection and in the younger age strata, consistent with our previous observations^7^ and with other COVID-19 vaccines.^16-19^ Notably, solicited adverse reactions were reported less frequently and were milder with the optimised formulations in the current study compared to the formulations tested in the previous Phase 1/2 trial.^7^ We observed a similar reactogenicity profile between naïve and non-naïve individuals, in contrast to reports from other vaccines which show higher rates of solicited reactions in seropositive vaccinees.^20,21^ Increased reactogenicity has been consistently noted for AS03-adjuvanted vaccines when compared with the corresponding unadjuvanted vaccines, for pandemic influenza vaccines^22-25^ and for CoV2 preS dTM formulations evaluated in our previous Phase 1/2 study.^7^ Of note, the proportions of participants with local and systemic reactogenicity (overall and grade 3) after two vaccine doses were higher in our study population than previously observed with AS03-adjuvanted pandemic influenza vaccines^23-25^ or in Phase I trials of the AS03-adjuvanted SARS-CoV-2 virus-like particle vaccine^19^ and the AS03-adjuvanted recombinant full-length S protein produced in CHO cells;^18^ however, they were similar to those observed in the pivotal clinical study of the SARS-CoV-2 mRNA-1273 vaccine after two vaccine doses.^26^ While our study did not include a placebo group which may impact the reporting of reactogenicity, these observations taken together suggest that the combination of the adjuvant and antigen contribute to the reactogenicity profile of candidate SARS-CoV-2 vaccines. Transient, self-resolving events of elevated blood pressure not associated with symptoms (except in one participant) were observed shortly after vaccination, which could be consistent with a procedure-related noradrenergic discharge around the time of vaccination.^27,28^

Almost all (≥97%) SARS-CoV-2 naïve participants attained a 4-fold rise in NAb titres to the D614G variant, regardless of age strata, presence of high-risk medical condition or antigen dose level. The magnitude of the NAb response observed at D36 in the naïve study population was comparable to that observed for a panel of human convalescent sera when measured using the same PsVN D614G assay, laboratory, and time frame. Amongst non-naïve adults in our study, a single injection increased D614G NAb titres to levels above those observed after two injections amongst naïve adults and exceeding those measured in the convalescent sera. Our findings are in line with the robust antibody titres previously observed after a single dose of the BNT162b2 (Pfizer-BioNTech) or mRNA-1273 (Moderna) SARS-CoV-2 vaccines in SARS-CoV-2 seropositive patients, which were also similar or greater in magnitude than achieved after two doses in naïve individuals.^29-32^ It is interesting to note the variability in response after the first dose in non-naïve participants suggesting that previous infection and priming is variable.

Although only limited information regarding correlates of protection is available,^33^ recent work has modelled the correlation between the ratio of NAb responses in vaccinees to convalescent sera and the observed vaccine efficacy to account for differences across assays and convalescent sera.^34,35^ In these models, ratios of 1 correlate to vaccine efficacy of 80–90% and ratios of 0·8 correlate to vaccine efficacy of 70–80%. In the current study, the ratio of NAb titres to convalescent sera among the younger naïve adults ranged between 1·38 and 2·40 across the groups, and among the older naïve adults the ratio was 0·65–0·81. However, this correlation was modelled for NAb responses and efficacy against homologous variants, or variants with small drifts. The lower responses to the Beta variant seen in this study, consistent with recent data from other authorised or investigational COVID vaccines,^36-39^ suggest NAb titres and predicted vaccine efficacy against heterologous variants are likely to be lower.^35^

Based on the interim safety and immunogenicity data described here, the CoV2 preS dTM-AS03 candidate vaccine has progressed to Phase 3 efficacy evaluation [NCT04904549]. Given that the reactogenicity and safety profiles were similar across antigen dose groups, the choice of antigen dose to progress to Phase 3 efficacy evaluation was largely dependent on the observed immunogenicity profile in naïve adults. Selection of the 10 μg total S antigen for a monovalent vaccine, over the 5 μg dose, is expected to mitigate the potential impact of variant circulation as it may provide higher cross-reactive antibody titres against variant strains in naïve individuals, albeit we did not observe a clear dose response relationship to the Beta variant in this study. Based on these models of predicted vaccine efficacy curves, we would expect any potential difference in vaccine protection between antigen dose groups to be limited. Furthermore, in the context of a pandemic, a lower antigen dose would translate into a significant increase in vaccine supply. In the Phase 3 study, a bivalent D614G + Beta variant AS03-adjuvanted vaccine with a 5 μg D614G component + 5 μg Beta component antigen dose is being evaluated. As the 5 μg dose in the naïve population in this study provided homologous NAb responses to the D614G variant comparable to convalescent sera, it is expected that a similar homologous response would be elicited by the Beta component of a bivalent vaccine.

Fractionation of doses has been suggested as an important strategy for meeting global vaccine demand,^40^ particularly in the context of booster vaccines. The robust NAb responses observed following a single injection of the 5 μg antigen dose formulation in SARS-CoV-2 non-naïve participants suggest that a single dose of 5 μg CoV2 preS dTM antigen with AS03 adjuvant may be sufficient for boosting previously primed individuals. This Phase 2 study has been amended to include cohorts of previously vaccinated individuals to evaluate a single 5 μg antigen dose as a booster vaccine.

The number of participants in this study limit the assessment of rare SAEs and AESIs, although continued follow-up and the large sample size recruited for the subsequent Phase 3 study, will provide a robust dataset for further safety evaluation. It should be noted that we have limited information on the donors of the convalescent sera used in this study; the clinical severity of COVID-19 ranged from mild to severe. Other limitations include that information on the durability of the immune response is not available from this interim analysis, and that antigen doses lower than 5 μg, which could be of interest for boosting primed individuals, were not evaluated here.

In summary, two doses of the CoV2 preS dTM-AS03 vaccine candidate demonstrated an acceptable safety profile and robust immunogenicity in SARS-CoV-2 naïve adults, including in individuals aged 60 years and older and those with high-risk medical conditions. Based on these results, two formulations of the CoV2 preS dTM-AS03 vaccine candidate, a monovalent D614G and a bivalent D614G and Beta variant vaccine, are undergoing efficacy evaluation in Phase 3 trials. Furthermore, the high neutralising titres and acceptable safety after a single vaccine dose observed in participants with evidence of prior SARS-CoV-2 infection indicate the possibility of developing a formulation with a lower antigen dose, and a single dose vaccination strategy, for use as a booster for previously primed adults.

## Data Availability

Qualified researchers can request access to patient-level data and related study documents, including the clinical study report, study protocol with any amendments, blank case report forms, statistical analysis plan, and dataset specifications. Patient-level data will be anonymised and study documents will be redacted to protect the privacy of trial participants. Further details on Sanofi's data sharing criteria, eligible studies, and process for requesting access can be found at https://www.clinicalstudydatarequest.com.

## Author Contributions

SS, MB, ALC, RC, BF, NG, YM, NGR, NM, LS, MHG, MKJ, MK, TT, JT, CAD and SS contributed to the concept or design of the study and data analysis and interpretation; HJ contributed to the conception or design of the study; SRW contributed to the concept or design of the study, data analysis and interpretation and data acquisition; GdB and JS were involved in data acquisition and data analysis and interpretation; AJ, AB, DD, BE, MK, DMR, SM, OO, VNR, RS, ER, NR, LDS, JW, DVB, RC, DL, SM, CM, SR, AS contributed to data acquisition; AC, NR, MAC and FTDS were involved in the analysis and interpretation of the data; RM was involved in conception or design of the study and data acquisition. All authors were involved in drafting or critically revising the manuscript, and all authors approved the final version and are accountable for the accuracy and integrity of the manuscript. All authors had full access to all the data and accept responsibility to submit for publication. At least two authors (AC and BF) have accessed and verified the data.

## Declaration of Interests

SS, MB, ALC, AC, RMC, BF, YM, ER, NR, DVB, GdB, RC, MHG, DL, HJ, RM, SM, CM, CAD and SSv are Sanofi Pasteur employees and may hold stock options. RMC reports owning shareholder and vesting options in Sanofi Pasteur. Additionally, SS, RMC, GdB, CAD and SSv, are inventors on a pending patent application filed by Sanofi Pasteur and GlaxoSmithKline for the development of the CoV-2 dTM vaccine. LS, MK, MAC, and FTDS are employed by the GlaxoSmithKline (GSK) group of companies and hold restricted shares in the GSK group of companies. AJ was contracted by Sanofi Pasteur and DD received funding from Sanofi Pasteur for his institution, George Washington University, to serve as a clinical site for the vaccine trial; and a percentage of university salary is allocated from this budget. NG reports receiving funding and study materials from Sanofi and funding from NIAID through the the COVID-19 Prevention Network (CoVPN) which provides research grants to her institution the Fred Hutchinson Cancer Research Center. MCK received NIAID funding support for the current study, paid to the clinical trials site for the COVID-19 prevention network and HIV vaccine trails network; Sanofi Pasteur contracted the site to conduct this clinical trial. DMR received a research grant from Sanofi Pasteur for this study. OO received a Gilead Sciences paid grant to Yale, and consulting fees from Gilead Sciences. OO also reports that Gilead, Integrity CE and Medscape paid for lectures, presentations, manuscript writing or educational events and that Gilead and VIIV paid for participation on a data safety monitoring or advisory board. VNR received funding from Sanofi for the CoVPN and Vaccine Treatment and Evaluation Units (VTEU) for COVID-19 vaccine clinical trials, and payments from Pfizer to her institution for the conduct of COVID-19 vaccine clinical trials. RS received clinical trial investigator site contracts with AES Synexus. NGR received a grant from Sanofi Pasteur to conduct this study and received grants from Sanofi, Merck, Pfizer, Quidel, Lilly in the past 36 months. NGR received a payment from IDAC and participated on an advisory board or safety monitoring for Micron, ICON and EMMES and reports having a leadership or fiduciary role (paid or unpaid) for IDSA, VTEU and ARLG. LDS received a research grant from Sanofi Pasteur and SRW and JW were contracted by Sanofi Pasteur. Authors report receiving funding from the US government through BARDA and the NIH for the clinical trial. AB, BE, NM, SM, MKJ, SR, JS, TT and JT declare no conflict of interest.

The views presented here are those of the authors and do not purport to represent those of the Department of the Army.

## Acknowledgements

The authors thank all participants, investigators, and study site personnel who took part in this study. The authors acknowledge Juliette Gray of inScience Communications, Springer Healthcare, London, UK, for providing editorial assistance with the preparation of this manuscript, funded by Sanofi Pasteur. The authors also thank Isabel Grégoire for providing editorial assistance and manuscript coordination on behalf of Sanofi Pasteur Funding was provided by Sanofi Pasteur and by federal funds from the Biomedical Advanced Research and Development Authority, part of the office of the Assistant Secretary for Preparedness and Response at the U.S. Department of Health and Human Services in collaboration with the U.S. Department of Defense Joint Program Executive Office for Chemical, Biological, Radiological and Nuclear Defense under Contract # W15QKN-16-9-1002. This work was done in collaboration with GlaxoSmithKline, who provided access to, and use of, the AS03 Adjuvant System.

## References

1. Jensen N, Kelly AH, Avendano M. The COVID-19 pandemic underscores the need for an equity-focused global health agenda. Humanities and Social Sciences Communications 2021; 8(1): 15.

2. World Bank. Global Economic Prospects, June 2021. Washington, DC: World Bank. doi:10.1596/978-1-4648-1665-9. 2021.

3. World Health Organization. The impact of COVID-19 on global health goals. Available at https://www.who.int/news-room/spotlight/the-impact-of-covid-19-on-global-health-goals. Accessed 06 June 2021.

4. Regulatory Affairs Professionals Society (RAPS). COVID-19 vaccine tracker. Available at https://www.raps.org/news-and-articles/news-articles/2020/3/covid-19-vaccine-tracker. Accessed on 27 April 2021. 2021.

5. The U.S. Food & Drug Administration. FDA NEWS RELEASE: FDA Approves First COVID-19 Vaccine. AVailable at https://www.fda.gov/news-events/press-announcements/fda-approves-first-covid-19-vaccine. Accessed 07 September 2021. 2021.

6. Kramer DB, Opel DJ, Parasidis E, Mello MM. Choices in a Crisis — Individual Preferences among SARS-CoV-2 Vaccines. New England Journal of Medicine 2021; 384(17): e62.

7. Goepfert PA, Fu B, Chabanon AL, et al. Safety and immunogenicity of SARS-CoV-2 recombinant protein vaccine formulations in healthy adults: interim results of a randomised, placebo-controlled, phase 1-2, dose-ranging study. Lancet Infect Dis 2021.

8. Arunachalam PS, Walls AC, Golden N, et al. Adjuvanting a subunit COVID-19 vaccine to induce protective immunity. Nature 2021; 594(7862): 253–8.

9. Centers for disease control and prevention. Underlying Medical Conditions Associated with High Risk for Severe COVID-19: Information for Healthcare Providers. Available at https://www.cdc.gov/coronavirus/2019-ncov/hcp/clinical-care/underlyingconditions.html. Accessed on 30 April 2021. 2021.

10. Garçon N, Vaughn DW, Didierlaurent AM. Development and evaluation of AS03, an Adjuvant System containing α-tocopherol and squalene in an oil-in-water emulsion. Expert review of vaccines 2012; 11(3): 349–66.

11. Petropoulos CJ, Parkin NT, Limoli KL, et al. A Novel Phenotypic Drug Susceptibility Assay for Human Immunodeficiency Virus Type 1. Antimicrobial Agents and Chemotherapy 2000; 44(4): 920–8.

12. Richman DD, Wrin T, Little SJ, Petropoulos CJ. Rapid evolution of the neutralizing antibody response to HIV type 1 infection. Proceedings of the National Academy of Sciences 2003; 100(7): 4144–9.

13. Whitcomb JM, Huang W, Fransen S, et al. Development and Characterization of a Novel Single-Cycle Recombinant-Virus Assay To Determine Human Immunodeficiency Virus Type 1 Coreceptor Tropism. Antimicrobial Agents and Chemotherapy 2007; 51(2): 566–75.

14. Tavares Da Silva F, De Keyser F, Lambert P-H, Robinson WH, Westhovens R, Sindic C. Optimal approaches to data collection and analysis of potential immune mediated disorders in clinical trials of new vaccines. Vaccine 2013; 31(14): 1870–6.

15. Newcombe RG. Two-sided confidence intervals for the single proportion: comparison of seven methods. Statistics in medicine 1998; 17(8): 857–72.

16. Heath PT, Galiza EP, Baxter DN, et al. Safety and Efficacy of NVX-CoV2373 Covid-19 Vaccine. New England Journal of Medicine 2021.

17. Mulligan MJ, Lyke KE, Kitchin N, et al. Phase I/II study of COVID-19 RNA vaccine BNT162b1 in adults. Nature 2020; 586(7830): 589–93.

18. Richmond P, Hatchuel L, Dong M, et al. Safety and immunogenicity of S-Trimer (SCB-2019), a protein subunit vaccine candidate for COVID-19 in healthy adults: a phase 1, randomised, double-blind, placebo-controlled trial. Lancet (London, England) 2021; 397(10275): 682–94.

19. Ward BJ, Gobeil P, Séguin A, et al. Phase 1 randomized trial of a plant-derived virus-like particle vaccine for COVID-19. Nature Medicine 2021; 27: 1071–8.

20. Krammer F, Srivastava K, Alshammary H, et al. Antibody Responses in Seropositive Persons after a Single Dose of SARS-CoV-2 mRNA Vaccine. New England Journal of Medicine 2021; 384(14): 1372–4.

21. Ebinger JE, Fert-Bober J, Printsev I, et al. Antibody responses to the BNT162b2 mRNA vaccine in individuals previously infected with SARS-CoV-2. Nat Med 2021; 27(6): 981–4.

22. Cohet C, van der Most R, Bauchau V, et al. Safety of AS03-adjuvanted influenza vaccines: A review of the evidence. Vaccine 2019; 37(23): 3006–21.

23. Jackson LA, Campbell JD, Frey SE, et al. Effect of Varying Doses of a Monovalent H7N9 Influenza Vaccine With and Without AS03 and MF59 Adjuvants on Immune Response: A Randomized Clinical Trial. JAMA 2015; 314(3): 237–46.

24. Levie K, Leroux-Roels I, Hoppenbrouwers K, et al. An adjuvanted, low-dose, pandemic influenza A (H5N1) vaccine candidate is safe, immunogenic, and induces cross-reactive immune responses in healthy adults. The Journal of infectious diseases 2008; 198(5): 642–9.

25. Vesikari T, Pepin S, Kusters I, Hoffenbach A, Denis M. Assessment of squalene adjuvanted and non-adjuvanted vaccines against pandemic H1N1 influenza in children 6 months to 17 years of age. Hum Vaccin Immunother 2012; 8(9): 1283–92.

26. Baden LR, El Sahly HM, Essink B, et al. Efficacy and Safety of the mRNA-1273 SARS-CoV-2 Vaccine. New England Journal of Medicine 2020; 384(5): 403–16.

27. World Health Organization. Immunization stress-related responses - a manual. Accessible at https://www.who.int/publications/i/item/978-92-4-151594-8. Accessed on 25 May 2021. 2019.

28. Meylan S, Livio F, Foerster M, Genoud PJ, Marguet F, Wuerzner G. Stage III Hypertension in Patients After mRNA-Based SARS-CoV-2 Vaccination. Hypertension 2021; 77(6): e56–e7.

29. Manisty C, Otter AD, Treibel TA, et al. Antibody response to first BNT162b2 dose in previously SARS-CoV-2-infected individuals. The Lancet 2021; 397(10279): 1057–8.

30. Prendecki M, Clarke C, Brown J, et al. Effect of previous SARS-CoV-2 infection on humoral and T-cell responses to single-dose BNT162b2 vaccine. The Lancet 2021; 397(10280): 1178–81.

31. Krammer F, Srivastava K, Simon V. Robust spike antibody responses and increased reactogenicity in seropositive individuals after a single dose of SARS-CoV-2 mRNA vaccine. medRxiv preprint server 2021: https://www.medrxiv.org/content/medrxiv/early/2021/02/01/.01.29.21250653.full.pdf. Accessed 06 June 2021.

32. Saadat S, Tehrani ZR, Logue J, et al. Single Dose Vaccination in Healthcare Workers Previously Infected with SARS-CoV-2. medRxiv preprint server 2021: https://www.medrxiv.org/content/medrxiv/early/2021/02/18/.01.30.21250843.full.pdf. Accessed 06 June 2021.

33. Gilbert PB, Montefiori DC, McDermott A, et al. Immune Correlates Analysis of the mRNA-1273 COVID-19 Vaccine Efficacy Trial. medRxiv 2021.

34. Earle KA, Ambrosino DM, Fiore-Gartland A, et al. Evidence for antibody as a protective correlate for COVID-19 vaccines. medRxiv preprint server: https://www.medrxiv.org/content/medrxiv/early/2021/03/20/.03.17.20200246.full.pdf. Accessed 06 June 2021.

35. Khoury DS, Cromer D, Reynaldi A, et al. What level of neutralising antibody protects from COVID-19? medRxiv preprint server: https://www.medrxiv.org/content/medrxiv/early/2021/03/11/.03.09.21252641.full.pdf. Accessed 06 June 2021.

36. Liu Y, Liu J, Xia H, et al. Neutralizing Activity of BNT162b2-Elicited Serum. New England Journal of Medicine 2021; 384(15): 1466–8.

37. Madhi SA, Baillie V, Cutland CL, et al. Efficacy of the ChAdOx1 nCoV-19 Covid-19 Vaccine against the B.1.351 Variant. New England Journal of Medicine 2021; 384(20): 1885–98.

38. Wang P, Nair MS, Liu L, et al. Antibody Resistance of SARS-CoV-2 Variants B.1.351 and B.1.1.7. bioRxiv preprint server 2021: https://www.biorxiv.org/content/biorxiv/early/2021/02/12/.01.25.428137.full.pdf. Accessed 06 June 2021.

39. Hoffmann M, Arora P, Groß R, et al. SARS-CoV-2 variants B.1.351 and P.1 escape from neutralizing antibodies. Cell 2021; 184(9): 2384–93.e12.

40. Cowling BJ, Lim WW, Cobey S. Fractionation of COVID-19 vaccine doses could extend limited supplies and reduce mortality. Nature Medicine 2021; 27(8): 1321–3.

