## Supplementary materials for "Safety and immunogenicity of a SARS-CoV-2 recombinant protein vaccine with AS03 adjuvant in healthy adults: interim findings from a phase 2, randomised, dose-finding, multi-centre study"

**Sridhar S *et al.***

**Contents**

List of VAT00002 COVID-19 study contributors	2

Methods 1. Participant inclusion/exclusion criteria.	3

Methods 2. Determination of SARS-CoV-2 naïve or non-naïve status for immunogenicity assessment	4

Table S1. High-risk medical conditions of study participants (FAS)	5

Table S2: Safety overview after any injection in all participants by age stratum regardless of SARS-CoV-2 positivity at baseline (D1), and for all ages by SARS-CoV-2 positivity at baseline (SafAS)	6

Table S3. Most frequently reported unsolicited adverse reactions (MedDRA preferred terms) (SafAS)	11

Table S4. Neutralising antibody response after two injections in naïve participants by presence of high-risk medical condition (PPAS-naïve D1+D22)	12

Table S5. Binding antibody profile, overall and by age, at D36 (PPAS-naïve D1+D22)	15

Figure S1. Participant flow through the interim study period (up to study day 43)	16

Figure S2. Solicited injection site and solicited systemic adverse reactions occurring over the first seven days after the second vaccination for younger adults (A; 18–59 years) and older adults (B; ≥60 years)	17

Figure S3. Neutralising antibody response to the Beta variant, 14 days after the second injection of CoV2 preS dTM-AS03 (all dose groups combined), by SARS-CoV-2 naïve status…………………………………………21
Figure S4. CMI analyses – ratios of fold-rises for IFN-γ (A), IL-2 (B) and TNF-α (C) to IL-4, IL-5 and IL-13 at Day 22 and Day 36 (CMIAS)…………………………………………………………………………………….23

**List of contributors**

**Brigham and Women’s Hospital, Boston, MA, USA:** Lindsey R. Baden, Jennifer A. Johnson, Amy C. Sherman, Michaël Desjardins, Jane A. Kleinjan, Jon A. Gothing, Megan P. Powell, Julia E. Klopfer; **Charles R. Drew University:** David Hardy, MD; Naureen Taureen, MD; **Demedica, San Pedro Sula, Honduras:** Luis Cousin MD; Delmy Mejía MD; Jorge Cortés MD ; Catherine Hardy MD; Engels Cardona MD; Allan R. Bueso MD; Blanca Panting MD; Marcelo Forgas MD; Belinda Andino MD; **Emory University School of Medicine, Atlanta, GA, USA:** Amy Anderson, Cecilia Losada, MD; Jessica Traenkner, PA; Nicholas Scanlon, MD with support from the NIH R38 Stimulating Access to Research in Residency (StARR) grant (5R38AI140299-02); **INVERIME investigators:** Silvia M Rivera MD, Sara E Rivera MD and Celim Chau MD; **New York University Grossman School of Medicine, New York, USA:** Tamia Davis, Heather Givans, Ramin Sedaghat Herati, Alexander McMeeking, Mark J. Mulligan, Mary Olson, Lalitha Parameswaran, Purvi Parikh, Rebecca Pellett Madan, Bo Shopsin, Janine Sullivan, Elizabeth Veneskey, Mahija Vucetovic; **The George Washington School of Medicine and Health Sciences, Washington, DC, USA:** Elissa Malkin DO, Caroline Thoreson PA-C, Marc Siegel MD; **Yale University School of Medicine, New Haven, Connecticut, USA:** Jessica Tuan MD.

### Methods 1. Participant inclusion/exclusion criteria.

Adults, aged 18 years and older, were eligible. Exclusion criteria included women who were pregnant or lactating, or, for those of childbearing potential, not using an effective method of contraception or abstinence from at least 4 weeks prior to the first vaccination until at least 12 weeks after the last vaccination; known systemic hypersensitivity to any of the vaccine components, or history of a life-threatening reaction to a vaccine containing any of the same substances; dementia or any other cognitive condition at a stage that could interfere with following the trial procedures based on Investigator or designee's judgment; self-reported thrombocytopenia, contraindicating intramuscular (IM) vaccination based on Investigator or designee's judgment; bleeding disorder, or receipt of anticoagulants in the past 21 days preceding inclusion, contraindicating IM vaccination based on Investigator or designee's judgment; unstable acute or chronic illness that in the opinion of the Investigator or designee poses additional risk as a result of participation or that could interfere with the trial procedures; receipt of solid-organ or bone marrow transplants in the past 180 days; receipt of anti-cancer chemotherapy in the last 90 days; moderate or severe acute illness/infection (according to investigator judgment) on the day of vaccination or febrile illness (temperature ≥38.0°C [≥100.4°F]). A prospective participant should not be included in the trial until the condition has resolved or the febrile event has subsided; receipt of any vaccine in the 30 days preceding or on the day of the first trial vaccination or planned receipt of any vaccine in the 30 days following the second trial vaccination except for influenza vaccination, which may be received at any time in relation to trial intervention; prior administration of a coronavirus vaccine (SARS-CoV-2, SARS-CoV, Middle East Respiratory Syndrome [MERS-CoV]); participation at the time of trial enrollment (or in the 30 days preceding the first trial vaccination) or planned participation during the present trial period in another clinical trial investigating a vaccine, drug, medical device, or medical procedure.

To allow evaluation of vaccine performance in high-risk groups, individuals with high-risk medical conditions (including those considered to potentially increase the risk for severe COVID-19 illness^9^) or those who were immunocompromised for other reasons, were not excluded from the study.

### Methods 2. Determination of SARS-CoV-2 naïve or non-naïve status for immunogenicity assessment

| **Status** | **S-ELECSYS** | **N-ELECSYS** | **NAAT** | **Test result** |
| --- | --- | --- | --- | --- |
| Naïve D1+D22 | Neg at D1 | Neg at D1 AND D22 | Neg at D1 AND D22 | All negative at D1 and D22 |
| Naïve D1 | Neg at D1 | Neg at D1 | Neg at D1 | All negative at D1 |
| Non-Naïve D1/D22 | Pos at D1 | Pos at D1 OR D22 | Pos at D1 OR D22 | Any positive result at D1or D22 |
| Non-Naïve D1 | Pos at D1 | Pos at D1 | Pos at D1 | Any positive result at D1 |

SARS-CoV-2 naïve or non-naïve status was determined for all participants on serological samples using ELECSYS electrochemiluminescence immunoassays for anti-SARS-CoV-2 anti-S (S-ELECSYS; Elecsys Anti-SARS-CoV-2 S, Roche, Indianapolis, IN, USA EUA202698) on D1 only and anti-nucleocapsid (N) (N-ELECSYS; Elecsys Anti-SARS-CoV-2, Roche, EUA200514) on D1 and D22, and on nasopharyngeal swabs using nucleic acid amplification tests (NAAT; Abbott RealTime SARS-CoV-2 assay, EUA200023) on D1 and D22, following the manufacturer’s instructions. SARS-CoV-2 naïve participants at D1 and D22 (naïve D1+D22) were those who tested negative for S-ELECSYS at D1 and negative for both N-ELECSYS and NAAT at both timepoints; the non-naïve subset of participants tested positive for at least one of the three tests at D1 and/or D22 (non-naïve D1/D22)

### Table S1. High-risk medical conditions of study participants (FAS)

|  | **Low-dose (N=240) n (%)** | **Medium-dose (N=239)  n (%)** | **High-dose (N=242)  n (%)** |
| --- | --- | --- | --- |
| At least one high-risk medical condition* | 151 (62·9) | 143 (59·8) | 143 (59·1) |
| Obesity^†^ | 80 (33·3) | 92 (38·5) | 87 (36·0) |
| Hypertension | 84 (35·0) | 80 (33·5) | 75 (31·0) |
| Type 2 diabetes | 32 (13·3) | 26 (10·9) | 28 (11·6) |
| Smoking | 21 (8·8) | 15 (6·3) | 19 (7·9) |
| Cancer | 15 (6·3) | 15 (6·3) | 12 (5·0) |
| Moderate-to-severe asthma | 8 (3·3) | 8 (3·3) | 13 (5·4) |
| Heart Conditions | 9 (3·8) | 5 (2·1) | 7 (2·9) |
| Coronary artery disease or cardiomyopathies | 8 (3·3) | 4 (1·7) | 6 (2·5) |
| Immunocompromised state from causes other than solid organ transplant | 4 (1·7) | 5 (2·1) | 6 (2·5) |
| Chronic obstructive pulmonary disease | 6 (2·5) | 2 (0·8) | 4 (1·7) |
| Hepatic disease | 4 (1·7) | 5 (2·1) | 2 (0·8) |
| Cerebrovascular disease | 2 (0·8) | 5 (2·1) | 2 (0·8) |
| Neurologic conditions | 1 (0·4) | 1 (0·4) | 4 (1·7) |
| Chronic kidney disease | 0 | 0 | 4 (1·7) |
| Cystic fibrosis | 2 (0·8) | 1 (0·4) | 0 |
| Immunocompromised state from solid organ transplant | 1 (0·4) | 0 | 1 (0·4) |
| Type 1 diabetes | 0 | 1 (0·4) | 1 (0·4) |
| Pulmonary fibrosis | 1 (0·4) | 0 | 0 |
| Thalassemia | 1 (0·4) | 0 | 0 |
| Sickle cell disease | 0 | 0 | 0 |

n: number of subjects fulfilling the item listed
*High-risk conditions are those considered to be associated with an increased risk of severe COVID-19, as detailed at https://www.cdc.gov/coronavirus/2019-ncov/science/science-briefs/underlying-evidence-table.html; ^†^Obesity: body mass index of 30 kg/m^2^ or higher

### Table S2: Safety overview after any injection in all participants by age stratum regardless of SARS-CoV-2 positivity at baseline (D1), and for all ages by SARS-CoV-2 positivity at baseline (SafAS)

|  | **Low-dose (N=240)** | | | **Medium-dose (N=240)** | | | **High-dose (N=241)** | | | |
| --- | --- | --- | --- | --- | --- | --- | --- | --- | --- | --- |
|  | **n/M** | **%** | **(95% CI)** | **n/M** | **%** | **(95% CI)** | **n/M** | | **%** | **(95% CI)** |
| **Total participants (SafAS)** |  |  |  |  |  |  |  | |  |  |
| **All ages** |  |  |  |  |  |  |  | |  |  |
| Immediate unsolicited AE | 2/240 | 0.8 | (0·1; 3·0) | 1/240 | 0·4 | (0; 2·3) | 1/241 | | 0·4 | (0; 2·3) |
| Immediate unsolicited AR | 1/240 | 0·4 | (0; 2·3) | 1/240 | 0·4 | (0; 2·3) | 0/241 | | 0 | (0; 1·5) |
| ***Up to 7 days after vaccination*** |  |  |  |  |  |  |  | |  |  |
| Any solicited reaction | 217/238 | 91·2 | (86·8; 94·5) | 213/237 | 89·9 | (85·3; 93·4) | 218/239 | | 91·2 | (86·9; 94·5) |
| Any grade 3 solicited reaction | 52/238 | 21·8 | (16·8 ; 27·6) | 49/237 | 20·7 | (15·7 ; 26·4) | 45/239 | | 18·8 | (14·1 ; 24·4) |
| Solicited injection site reaction | 201/238 | 84·5 | (79·2; 88·8) | 196/237 | 82·7 | (77·3; 87·3) | 200/239 | | 83·7 | (78·4; 88·1) |
| Grade 3 injection site reaction | 20/238 | 8·4 | (5·2 ; 12·7) | 23/237 | 9·7 | (6·3 ; 14·2) | 17/239 | | 7·1 | (4·2 ; 11·1) |
| Solicited systemic reaction | 191/238 | 80·3 | (74·6; 85·1) | 192/237 | 81·0 | (75·4; 85·8) | 185/239 | | 77·4 | (71·6; 82·5) |
| Grade 3 systemic reaction | 45/238 | 18·9 | (14·1 ; 24·5) | 41/237 | 17·3 | (12·7 ; 22·7) | 41/239 | | 17·2 | (12·6 ; 22·5) |
| ***Up to 21 days after vaccination*** |  |  |  |  |  |  |  | |  |  |
| Any unsolicited AE | 87/240 | 36·3 | (30·2; 42·7) | 79/240 | 32·9 | (27·0; 39·3) | 89/241 | | 36·9 | (30·8; 43·4) |
| Any unsolicited non-serious AE | 87/240 | 36·3 | (30·2 ; 42·7) | 79/240 | 32·9 | (27·0; 39·3) | 87/241 | | 36·1 | (30·0; 42·5) |
| Any grade 3 unsolicited AE | 5/240 | 2·1 | (0·7 ; 4·8) | 6/240 | 2·5 | (0·9; 5·4) | 19/241 | | 7·9 | (4·8; 12·0) |
| Any unsolicited AR | 47/240 | 19·6 | (14·8; 25·2) | 43/240 | 17·9 | (13·3; 23·4) | 35/241 | | 14·5 | (10·3; 19·6) |
| Any unsolicited non-serious AR | 47/240 | 19·6 | (14·8 ; 25·2) | 43/240 | 17·9 | (13·3; 23·4) | 35/241 | | 14·5 | (10·3; 19·6) |
| Any grade 3 unsolicited AR | 1/240 | 0·4 | (0 ; 2·3) | 3/240 | 1·3 | (0·3; 3·6) | 5/241 | | 2·1 | (0·7 ; 4·8) |
| Unsolicited non-serious injection site AR | 13/240 | 5·4 | (2·9; 9·1) | 18/240 | 7·5 | (4·5; 11·6) | 9/241 | | 3·7 | (1·7; 7·0) |
| Unsolicited non-serious systemic AE | 80/240 | 33·3 | (27·4; 39·7) | 67/240 | 27·9 | (22·3; 34·1) | 83/241 | | 34·4 | (28·5; 40·8) |
| Unsolicited non-serious systemic AR | 38/240 | 15·8 | (11·5; 21·1) | 29/240 | 12·1 | (8·2; 16·9) | 27/241 | | 11·2 | (7·5; 15·9) |
| ***Up to D43*** |  |  |  |  |  |  |  | |  |  |
| AE leading to study discontinuation | 0/240 | 0 | (0; 1·5) | 0/240 | 0 | (0; 1·5) | 0/241 | | 0 | (0; 1·5) |
| SAE | 0/240 | 0 | (0; 1·5) | 2/240 | 0·8 | (0·1; 3·0) | 2/241 | | 0·8 | (0·1; 3·0) |
| Grade 3 SAE | 0/240 | 0 | (0; 1·5) | 1/240 | 0·4 | (0; 2·3) | 1/241 | | 0·4 | (0; 2·3) |
| Related SAE | 0/240 | 0 | (0; 1·5) | 0/240 | 0 | (0; 1·5) | 0/241 | | 0 | (0; 1·5) |
| Death | 0/240 | 0 | (0; 1·5) | 0/240 | 0 | (0; 1·5) | 0/241 | | 0 | (0; 1·5) |
| AESI | 0/240 | 0 | (0; 1·5) | 0/240 | 0 | (0; 1·5) | 0/241 | | 0 | (0; 1·5) |
| MAAE | 20/240 | 8·3 | (5·2; 12·6) | 11/240 | 4·6 | (2·3; 8·1) | 31/241 | | 12·9 | (8·9; 17·8) |
| **18-59 years** |  |  |  |  |  |  |  | |  |  |
| Immediate unsolicited AE | 2/121 | 1·7 | (0·2 ; 5·8) | 1/120 | 0·8 | (0 ; 4·6) | 0/119 | | 0 | (0 ; 3·1) |
| Immediate unsolicited AR | 1/121 | 0·8 | (0 ; 4·5) | 1/120 | 0·8 | (0 ; 4·6) | 0/119 | | 0 | (0 ; 3·1) |
| ***Up to 7 days after vaccination*** |  |  |  |  |  |  |  | |  |  |
| Any solicited reaction | 111/119 | 93·3 | (87·2; 97·1) | 110/119 | 92·4 | (86·1; 96·5) | 112/117 | | 95·7 | (90·3; 98·6) |
| Any grade 3 solicited reaction | 35/119 | 29·4 | (21·4; 38·5) | 34/119 | 28·6 | (20·7; 37·6) | 28/117 | | 23·9 | (16·5; 32·7) |
| Solicited injection site reaction | 107/119 | 89·9 | (83·0; 94·7) | 104/119 | 87·4 | (80·1; 92·8) | 109/117 | | 93·2 | (87·0; 97·0) |
| Grade 3 injection site reaction | 16/119 | 13·4 | (7·9; 20·9) | 16/119 | 13·4 | (7·9; 20·9) | 11/117 | | 9·4 | (4·8; 16·2) |
| Solicited systemic reaction | 105/119 | 88·2 | (81·0; 93·4) | 103/119 | 86·6 | (79·1; 92·1) | 94/117 | | 80·3 | (72·0; 87·1) |
| Grade 3 systemic reaction | 31/119 | 26·1 | (18·4; 34·9) | 30/119 | 25·2 | (17·7; 34·0) | 24/117 | | 20·5 | (13·6; 29·0) |
| ***Up to 21 days after vaccination*** |  |  |  |  |  |  |  | |  |  |
| Any unsolicited AE | 50/121 | 41·3 | (32·4; 50·6) | 44/120 | 36·7 | (28·1; 45·9) | 44/119 | | 37·0 | (28·3; 46·3) |
| Any unsolicited non-serious AE | 50/121 | 41·3 | (32·4; 50·6) | 44/120 | 36·7 | (28·1; 45·9) | 43/119 | | 36·1 | (27·5; 45·4) |
| Any grade 3 unsolicited AE | 1/121 | 0·8 | (0; 4·5) | 3/120 | 2·5 | (0·5; 7·1) | 9/119 | | 7·6 | (3·5; 13·9) |
| Any unsolicited AR | 30/121 | 24·8 | (17·4; 33·5) | 27/120 | 22·5 | (15·4; 31·0) | 17/119 | | 14·3 | (8·5; 21·9) |
| Any unsolicited non-serious AR | 30/121 | 24·8 | (17·4; 33·5) | 27/120 | 22·5 | (15·4; 31·0) | 17/119 | | 14·3 | (8·5; 21·9) |
| Any grade 3 unsolicited AR | 0/121 | 0 | (0; 3·0) | 2/120 | 1·7 | (0·2; 5·9) | 1/119 | | 0·8 | (0; 4·6) |
| Unsolicited non-serious injection site AR | 7/121 | 5·8 | (2·4; 11·6) | 9/120 | 7·5 | (3·5; 13·8) | 5/119 | | 4·2 | (1·4; 9·5) |
| Unsolicited non-serious systemic AE | 46/121 | 38·0 | (29·3; 47·3) | 39/120 | 32·5 | (24·2; 41·7) | 42/119 | | 35·3 | (26·8; 44·6) |
| Unsolicited non-serious systemic AR | 25/121 | 20·7 | (13·8; 29·0) | 21/120 | 17·5 | (11·2; 25·5) | 12/119 | | 10·1 | (5·3; 17·8) |
| ***Up to D43*** |  |  |  |  |  |  |  | |  |  |
| AE leading to study discontinuation | 0/121 | 0 | (0 ; 3·0) | 0/120 | 0 | (0 ; 3·0) | 0/119 | | 0 | (0 ; 3·1) |
| SAE | 0/121 | 0 | (0 ; 3·0) | 1/120 | 0·8 | (0 ; 4·6) | 1/119 | | 0·8 | (0 ; 4·6) |
| Grade 3 SAE | 0/121 | 0 | (0 ; 3·0) | 0/120 | 0 | (0 ; 3·0) | 0/119 | | 0 | (0 ; 3·1) |
| Related SAE | 0/121 | 0 | (0 ; 3·0) | 0/120 | 0 | (0 ; 3·0) | 0/119 | | 0 | (0 ; 3·1) |
| Death | 0/121 | 0 | (0 ; 3·0) | 0/120 | 0 | (0 ; 3·0) | 0/119 | | 0 | (0 ; 3·1) |
| AESI | 0/121 | 0 | (0 ; 3·0) | 0/120 | 0 | (0 ; 3·0) | 0/119 | | 0 | (0 ; 3·1) |
| MAAE | 9/121 | 7·4 | (3·5 ; 13·7) | 5/120 | 4·2 | (1·4; 9·5) | 15/119 | | 12·6 | (7·2; 19·9) |
| **≥60 years** |  |  |  |  |  |  |  | |  |  |
| Immediate unsolicited AE | 0/119 | 0 | (0 ; 3·1) | 0/120 | 0 | (0 ; 3·0) | 1/122 | | 0·8 | (0 ; 4·5) |
| Immediate unsolicited AR | 0/119 | 0 | (0 ; 3·1) | 0/120 | 0 | (0 ; 3·0) | 0/122 | | 0 | (0 ; 3·0) |
| ***Up to 7 days after vaccination*** |  |  |  |  |  |  |  | |  |  |
| Any solicited reaction | 106/119 | 89·1 | (82·0 ; 94·1) | 103/118 | 87·3 | (79·9 ; 92·7) | 106/122 | | 86·9 | (79·6; 92·3) |
| Any grade 3 solicited reaction | 17/119 | 14·3 | (8·5 ; 21·9) | 15/118 | 12·7 | (7·3 ; 20·1) | 17/122 | | 13·9 | (8·3; 21·4) |
| Solicited injection site reaction | 94/119 | 79·0 | (70·6 ; 85·9) | 92/118 | 78·0 | (69·4 ; 85·1) | 91/122 | | 74·6 | (65·9; 82·0) |
| Grade 3 injection site reaction | 4/119 | 3·4 | (0·9 ; 8·4) | 7/118 | 5·9 | (2·4 ; 11·8) | 6/122 | | 4·9 | (1·8; 10·4) |
| Solicited systemic reaction | 86/119 | 72·3 | (63·3 ; 80·1) | 89/118 | 75·4 | (66·6 ; 82·9) | 91/122 | | 74·6 | (65·9; 82·0) |
| Grade 3 systemic reaction | 14/119 | 11·8 | (6·6 ; 19·0) | 11/118 | 9·3 | (4·7 ; 16·1) | 17/122 | | 13·9 | (8·3; 21·4) |
| ***Up to 21 days after vaccination*** |  |  |  |  |  |  |  | |  |  |
| Any unsolicited AE | 37/119 | 31·1 | (22·9 ; 40·2) | 35/120 | 29·2 | (21·2; 38·2) | 45/122 | | 36·9 | (28·3; 46·1) |
| Any unsolicited non-serious AE | 37/119 | 31·1 | (22·9 ; 40·2) | 35/120 | 29·2 | (21·2; 38·2) | 44/122 | | 36·1 | (27·6; 45·3) |
| Any grade 3 unsolicited AE | 4/119 | 3·4 | (0·9 ; 8·4) | 3/120 | 2·5 | (0·5; 7·1) | 10/122 | | 8·2 | (4·0; 14·6) |
| Any unsolicited AR | 17/119 | 14·3 | (8·5 ; 21·9) | 16/120 | 13·3 | (7·8; 20·7) | 18/122 | | 14·8 | (9·0 ; 22·3) |
| Any unsolicited non-serious AR | 17/119 | 14·3 | (8·5 ; 21·9) | 16/120 | 13·3 | (7·8; 20·7) | 18/122 | | 14·8 | (9·0 ; 22·3) |
| Any grade 3 unsolicited AR | 1/119 | 0·8 | (0 ; 4·6) | 1/120 | 0·8 | (0; 4·0) | 4/122 | | 3·3 | (0·9 ; 8·2) |
| Unsolicited injection site AR | 6/119 | 5·0 | (1·9 ; 10·7) | 9/120 | 7·5 | (3·5; 13·8) | 4/122 | | 3·3 | (0·9 ; 8·2) |
| Unsolicited systemic AE | 34/119 | 28·6 | (20·7 ; 37·6) | 28/120 | 23·3 | (16·1; 312·9) | 41/122 | | 33·6 | (25·3; 42·7) |
| Unsolicited systemic AR | 13/119 | 10·9 | (5·9 ; 18·0) | 8/120 | 6·7 | (2·9; 12·7) | 15/122 | | 12·3 | (7·0 ; 19·5) |
| ***Up to D43*** |  |  |  |  |  |  |  | |  |  |
| AE leading to study discontinuation | 0/119 | 0 | (0 ; 3·1) | 0/120 | 0 | (0 ; 3·0) | 0/122 | | 0 | (0 ; 3·0) |
| SAE | 0/119 | 0 | (0 ; 3·1) | 1/120 | 0·8 | (0 ; 4·6) | 1/122 | | 0·8 | (0 ; 4·5) |
| Grade 3 SAE | 0/119 | 0 | (0 ; 3·1) | 1/120 | 0·8 | (0 ; 4·6) | 1/122 | | 0·8 | (0 ; 4·5) |
| Related SAE | 0/119 | 0 | (0 ; 3·1) | 0/120 | 0 | (0 ; 3·0) | 0/122 | | 0 | (0 ; 3·0) |
| Death | 0/119 | 0 | (0 ; 3·1) | 0/120 | 0 | (0 ; 3·0) | 0/122 | | 0 | (0 ; 3·0) |
| AESI | 0/119 | 0 | (0 ; 3·1) | 0/120 | 0 | (0 ; 3·0) | 0/122 | | 0 | (0 ; 3·0) |
| MAAE | 11/119 | 9·2 | (4·7 ; 15·9) | 6/120 | 5·0 | (1·9 ; 10·6) | 16/122 | | 13·1 | (7·7; 20·45) |
| **All ages, SARS-CoV-2 naïve at baseline (D1)*** | | | | | | | | | | |
| Immediate unsolicited AE | 2/211 | 0·9 | (0·1 ; 3·4) | 1/210 | 0·5 | (0 ; 2·6) | 1/207 | 0·5 | | (0 ; 2·7) |
| Immediate unsolicited AR | 1/211 | 0·5 | (0 ;2·6) | 1/210 | 0·5 | (0 ; 2·6) | 0/207 | 0 | | (0 ;1·8) |
| ***Up to 7 days after vaccination*** |  |  |  |  |  |  |  |  | |  |
| Any solicited reactions | 191/209 | 91·4 | (86·7; 94·8) | 189/208 | 90·9 | (86·1 ; 94·4) | 189/206 | 91·7 | | (87·1; 95·1) |
| Solicited injection site reaction | 177/209 | 84·7 | (79·1; 89·3) | 175/208 | 84·1 | (78·4 ; 88·8) | 173/206 | 84·0 | | (78·2; 88·7) |
| Solicited systemic reaction | 171/209 | 81·8 | (75·9; 86·8) | 170/208 | 81·7 | (75·8 ; 86·7) | 161/206 | 78·2 | | (71·9; 83·6) |
| ***Up to 21 days after vaccination*** |  |  |  |  |  |  |  |  | |  |
| Any unsolicited AEs | 81/211 | 38·4 | (31·8; 45·3) | 69/210 | 32·9 | (26·5 ; 39·7) | 81/207 | 39·1 | | (32·4; 46·1) |
| Any unsolicited ARs | 45/211 | 21·3 | (16·0; 27·5) | 37/210 | 17·6 | (12·7 ; 23·5) | 34/207 | 16·4 | | (11·7; 22·2) |
| Any unsolicited non-serious AE | 81/211 | 38·4 | (31·8; 45·3) | 69/210 | 32·9 | (26·5 ; 39·7) | 79/207 | 38·2 | | (31·5; 45·2) |
| Any unsolicited non-serious AR | 45/211 | 21·3 | (16·0; 27·5) | 37/210 | 17·6 | (12·7 ; 23·5) | 34/207 | 16·4 | | (11·7; 22·2) |
| Unsolicited non-serious injection site AR | 12/211 | 5·7 | (3·0; 9·7) | 16/210 | 7·6 | (4·4 ; 12·1) | 9/207 | 4·3 | | (2·0; 8·1) |
| Unsolicited non-serious systemic AE | 75/211 | 35·5 | (29·1; 42·4) | 58/210 | 27·6 | (21·7 ; 34·2) | 75/207 | 36·2 | | (29·7; 43·2) |
| Unsolicited non-serious systemic AR | 37/211 | 17·5 | (12·7 ; 23·4) | 25/210 | 11·9 | (7·9 ; 17·1) | 26/207 | 12·6 | | (8·4 ; 17·9) |
| ***Up to D43*** |  |  |  |  |  |  |  |  | |  |
| AE leading to study discontinuation | 0/211 | 0 | (0 ; 1·7) | 0/210 | 0 | (0 ; 1·7) | 0/207 | 0 | | (0 ; 1·8) |
| SAE | 0/211 | 0 | (0 ; 1·7) | 1/210 | 0·5 | (0 ; 2·6) | 2/207 | 1·0 | | (0·1 ; 3·4) |
| Related SAE | 0/211 | 0 | (0 ; 1·7) | 0/210 | 0 | (0 ; 1·7) | 0/207 | 0 | | (0 ; 1·8) |
| Death | 0/211 | 0 | (0 ; 1·7) | 0/210 | 0 | (0 ; 1·7) | 0/207 | 0 | | (0 ; 1·8) |
| AESI | 0/211 | 0 | (0 ; 1·7) | 0/210 | 0 | (0 ; 1·7) | 0/207 | 0 | | (0 ; 1·8) |
| MAAE | 20/211 | 9·5 | (5·9 ; 14·3) | 9/210 | 4·3 | (2·0 ; 8·0) | 28/207 | 13·5 | | (9·2 ; 19·0) |
| **All ages, SARS-CoV-2 non-naïve-D01†** | | | | | | | | | | |
| Immediate unsolicited AE | 0/26 | 0 | (0 ; 13·2) | 0/28 | 0 | (0 ; 12·3) | 0/31 | 0 | | (0 ; 11·2) |
| Immediate unsolicited AR | 0/26 | 0 | (0 ; 13·2) | 0/28 | 0 | (0 ; 12·3) | 0/31 | 0 | | (0 ; 11·2) |
| ***Up to 7 days after vaccination*** |  |  |  |  |  |  |  |  | |  |
| Any solicited reaction | 23/26 | 88·5 | (69·8 ; 97·6) | 22/27 | 81·5 | (61·9 ; 93·7) | 27/30 | 90·0 | | (73·5 ; 97·9) |
| Solicited injection site reaction | 21/26 | 80·8 | (60·6 ; 93·4) | 20/27 | 74·1 | (53·7 ; 88·9) | 25/30 | 83·3 | | (65·3 ; 94·4) |
| Solicited systemic reaction | 17/26 | 65·4 | (44·3 ; 82·8) | 20/27 | 74·1 | (53·7 ; 88·9) | 22/30 | 73·3 | | (54·1 ; 87·7) |
| ***Up to 21 days after vaccination*** |  |  |  |  |  |  |  |  | |  |
| Any unsolicited AE | 4/26 | 15·4 | (4·4 ; 34·9) | 10/28 | 35·7 | (18·6 ; 55·9) | 8/31 | 25·8 | | (11·9 ; 44·6) |
| Any unsolicited AR | 1/26 | 3·8 | (0·1 ; 19·6) | 6/28 | 21·4 | (8·3 ; 41·0) | 1/31 | 3·2 | | (0·1 ; 16·7) |
| Any unsolicited non-serious AE | 4/26 | 15·4 | (4·4 ; 34·9) | 10/28 | 35·7 | (18·6 ; 55·9) | 8/31 | 25·8 | | (11·9 ; 44·6) |
| Any unsolicited non-serious AR | 1/26 | 3·8 | (0·1 ; 19·6) | 6/28 | 21·4 | (8·3 ; 41·0) | 1/31 | 3·2 | | (0·1 ; 16·7) |
| Unsolicited non-serious injection site AR | 1/26 | 3·8 | (0·1 ; 19·6) | 2/28 | 7·1 | (0·9 ; 23·5) | 0/31 | 0 | | (0 ; 11·2) |
| Unsolicited non-serious systemic AE | 3/26 | 11·5 | (2·4 ; 30·2) | 9/28 | 32·1 | (15·9 ; 52·4) | 8/31 | 25·8 | | (11·9 ; 44·6) |
| Unsolicited non-serious systemic AR | 0/26 | 0 | (0 ; 13·2) | 4/28 | 14·3 | (4·0 ; 32·7) | 1/31 | 3·2 | | (0·1 ; 16·7) |
| ***Up to D43*** |  |  |  |  |  |  |  |  | |  |
| AE leading to study discontinuation | 0/26 | 0 | (0 ; 13·2) | 0/28 | 0 | (0 ; 12·3) | 0/31 | 0 | | (0 ; 11·2) |
| SAE | 0/26 | 0 | (0 ; 13·2) | 1/28 | 3·6 | (0·1 ; 18·3) | 0/31 | 0 | | (0 ; 11·2) |
| Related SAE | 0/26 | 0 | (0 ; 13·2) | 0/28 | 0 | (0 ; 12·3) | 0/31 | 0 | | (0 ; 11·2) |
| Death | 0/26 | 0 | (0 ; 13·2) | 0/28 | 0 | (0 ; 12·3) | 0/31 | 0 | | (0 ; 11·2) |
| AESI | 0/26 | 0 | (0 ; 13·2) | 0/28 | 0 | (0 ; 12·3) | 0/31 | 0 | | (0 ; 11·2) |
| MAAE | 0/26 | 0 | (0 ; 13·2) | 2/28 | 7·1 | (0·9 ; 23·5) | 3/31 | 9·7 | | (2·0 ; 25·8) |

*SARS-CoV-2 naïve at D1 i.e. tested negative for S-ELECSYS, N-ELECSYS, and NAAT on D1; ^†^SARS-CoV-2 non-naïve at D1, i.e. tested positive for 1 or more of the tests on D1

AE = adverse event. AESI = adverse event of special interest. AR = adverse reaction. n = number of participants experiencing the endpoint. M = number of participants with available data for the relevant endpoint. MAAE = medically-attended adverse event. SAE = serious adverse event.

Related SAEs: reported by an investigator as related to the vaccine. SAEs for which information on the relationship was missing, were considered as related

### Table S3. Most frequently reported unsolicited adverse reactions (MedDRA preferred terms) (SafAS)

|  | **Low-dose (N=240)** | | **Medium-dose (N=240)** | | **High-dose (N=241)** | |
| --- | --- | --- | --- | --- | --- | --- |
|  | **n/M** | **n ARs** | **n/M** | **n ARs** | **n/M** | **n ARs** |
| **Fatigue** | 17/240 | 18 | 8/240 | 10 | 13/241 | 15 |
| **Nausea** | 7/240 | 7 | 5/240 | 5 | 2/241 | 2 |
| **Pruritus** | 0/240 | 0 | 4/240 | 5 | 0/241 | 0 |
| **Diarrhoea** | 3/240 | 3 | 2/240 | 2 | 1/241 | 1 |
| **Dizziness** | 2/240 | 2 | 2/240 | 2 | 3/241 | 3 |
| **Injection site rash** | 1/240 | 1 | 2/240 | 2 | 1/241 | 1 |
| **Decreased appetite** | 1/240 | 1 | 2/240 | 2 | 0/241 | 0 |
| **Lymphadenopathy** | 2/240 | 3 | 1/240 | 1 | 3/241 | 3 |

n = number of participants reporting the specified event. M = number of participants with available data for the relevant endpoint. n ARs = number of adverse reactions. SafAS = safety analysis set.

Adverse reactions were recorded using MedDRA preferred terms.

### Table S4. D614G binding antibody profile at D36, overall, by age and by SARS-CoV-2 naïve status (PPAS)

|  | **Low-dose (5 µg) (N=201)** | | **Medium-dose (10 µg) (N=207)** | | **High-dose (15 µg) (N=203)** | |
| --- | --- | --- | --- | --- | --- | --- |
| **Endpoint** | **n/M or M** | **% or GMC/GMCR**  **(95% CI)** | **n/M or M** | **% or GMC/GMCR**  **(95% CI)** | **n/M or M** | **% or GMC/GMCR**  **(95% CI)** |
| **PPAS-naïve D1+D22** | | | | | | |
| **All ages** | | | | | | |
| ≥2-fold rise (responders)* | 153/153 | 100 (97·6, 100) | 164/165 | 99·4 (96·7, 100) | 158/159 | 99·4 (96·5, 100) |
| ≥4-fold rise | 161/162 | 99·4 (96·6, 100) | 170/171 | 99·4 (96·8, 100) | 170/171 | 99·4 (96·8, 100) |
| GMCs | 168 | 15503 (13040, 18431) | 177 | 17188 (14560, 20289) | 176 | 17872 (14957, 21355) |
| GMCR | 162 | 1463 (1224, 1748) | 171 | 1692 (1422, 2014) | 171 | 1733 (1437, 2090) |
| **18–59 years** |  |  |  |  |  |  |
| ≥2-fold rise (responders)* | 74/74 | 100 (95·1, 100) | 75/75 | 100 (95·2, 100) | 78/78 | 100 (95·4, 100) |
| ≥4-fold rise | 79/79 | 100 (95·4, 100) | 78/78 | 100 (95·4, 100) | 81/81 | 100 (95·5, 100) |
| GMCs | 83 | 21739 (18095; 26116) | 82 | 25448 (20980, 30867) | 81 | 26549 (22179, 31779) |
| GMCR | 79 | 1950 (1595, 2384) | 78 | 2427 (1953, 3016) | 81 | 2684 (2231, 3230) |
| **≥60 years** |  |  |  |  |  |  |
| ≥2-fold rise (responders)* | 79/79 | 100 (95·4, 100) | 89/90 | 98·9 (94·0, 100) | 80/81 | 98·8 (93·3, 100) |
| ≥4-fold rise | 82/83 | 98·8 (93·5, 100) | 92/93 | 98·9 (94·2; 100) | 89/90 | 98·9 (94·0, 100) |
| GMCs | 85 | 11144 (8446, 14705) | 95 | 12249 (9604, 15623) | 95 | 12753 (9664, 16830) |
| GMCR | 83 | 1112 (839, 1475) | 93 | 1251 (974, 1606) | 90 | 1169 (870, 1570) |
| **PPAS non-naïve D1/D22** | | | | | | |
| **All ages** |  |  |  |  |  |  |
| ≥2-fold rise | 28/28 | 100 (87.7, 100) | 24/26 | 92.3 (74.9, 99.1) | 23/23 | 100 (85.2, 100) |
| ≥4-fold rise (responders)* | 28/28 | 100 (87.7, 100) | 24/26 | 92.3 (74.9, 99.1) | 23/23 | 100 (85.2, 100) |
| GMCs | 28 | 27565 (19544, 38879) | 26 | 26293 (17728, 38996) | 23 | 48699 (31012, 76475) |
| GMCR | 28 | 131 (56.4, 303) | 26 | 111 (45.5, 271) | 23 | 284 (134, 603) |
| **18–59 years** |  |  |  |  |  |  |
| ≥2-fold rise | 13/13 | 100 (75.3, 100) | 14/16 | 87.5 (61.7, 98.4) | 14/14 | 100 (76.8, 100) |
| >4-fold rise (responders)* | 13/13 | 100 (75.3, 100) | 14/16 | 87.5 (61.7, 98.4) | 14/14 | 100 (76.8, 100) |
| GMCs | 13 | 34939 (20779, 58748) | 16 | 32578 (21327, 49766) | 14 | 50322 (25193, 101 000) |
| GMCR | 13 | 195 (51.9, 732) | 16 | 96.3 (27.8, 334) | 14 | 371 (179, 769) |
| **≥60 years** |  |  |  |  |  |  |
| ≥2-fold rise | 15/15 | 100 (78.2, 100) | 10/10 | 100 (69.2, 100) | 9/9 | 100 (66.4, 100) |
| ≥4-fold rise (responders)* | 15/15 | 100 (78.2, 100) | 10/10 | 100 (69.2, 100) | 9/9 | 100 (66.4, 100) |
| GMCs | 15 | 22446 (13739, 36671) | 10 | 18660 (8054, 43229) | 9 | 46277 (24818, 86291) |
| GMCR | 15 | 92.4 (27.9 ; 306) | 10 | 139 (31.3 ; 621) | 9 | 187 (31.2 ; 1124) |

CI = confidence interval. GMC = geometric mean concentration. GMCR = geometric mean concentration ratio. LLOQ = lower limit of quantification. PPAS = per protocol analysis set.

*Responders were defined in the naive participant subset as those who had ≥2-fold rise in antibody titres from baseline to D36 and in the non-naïve subset as those who had ≥4-fold rise in antibody titres from baseline to D36.

### Table S5. D614G neutralising antibody response after two injections in naïve participants by presence of high-risk medical condition (PPAS-naïve D1+D22)

|  | **Low-dose N=168** | | **Medium-dose N=177** | | **High-dose N=176** | |
| --- | --- | --- | --- | --- | --- | --- |
|  | **M** | **GMT (95% CI)** | **M** | **GMT (95% CI)** | **M** | **GMT (95% CI)** |
| **Overall** | 165 | 2189 (1744, 2746) | 173 | 2269 (1792, 2873) | 172 | 2895 (2294, 3654) |
| ***With a high-risk medical condition*** | | | | | | |
| **All ages** | 99 | 2085 (1514, 2872) | 104 | 2023 (1480, 2764) | 98 | 2033 (1490, 2775) |
| **18-59 years** | 34 | 3241 (2127, 4938) | 37 | 3830 (2234, 6566) | 32 | 4678 (2903, 7537) |
| **≥60 years** | 65 | 1655 (1074, 2550) | 67 | 1422 (987, 2048) | 66 | 1358 (939, 1962) |
| ***Without a high-risk medical condition*** | | | | | | |
| **All ages** | 66 | 2354 (1725, 3213) | 69 | 2699 (1875, 3884) | 74 | 4622 (3323, 6428) |
| **18-59 years** | 48 | 2766 (1956, 3912) | 44 | 4056 (2680, 6136) | 49 | 5469 (3651, 8193) |
| **≥60 years** | 18 | 1532 (770, 3045) | 25 | 1317 (700, 2478) | 25 | 3323 (1844, 5987) |

### GMT = geometric mean titre. PPAS = per protocol analysis set.

### Figure S1. Participant flow through the interim study period (up to study day 43)

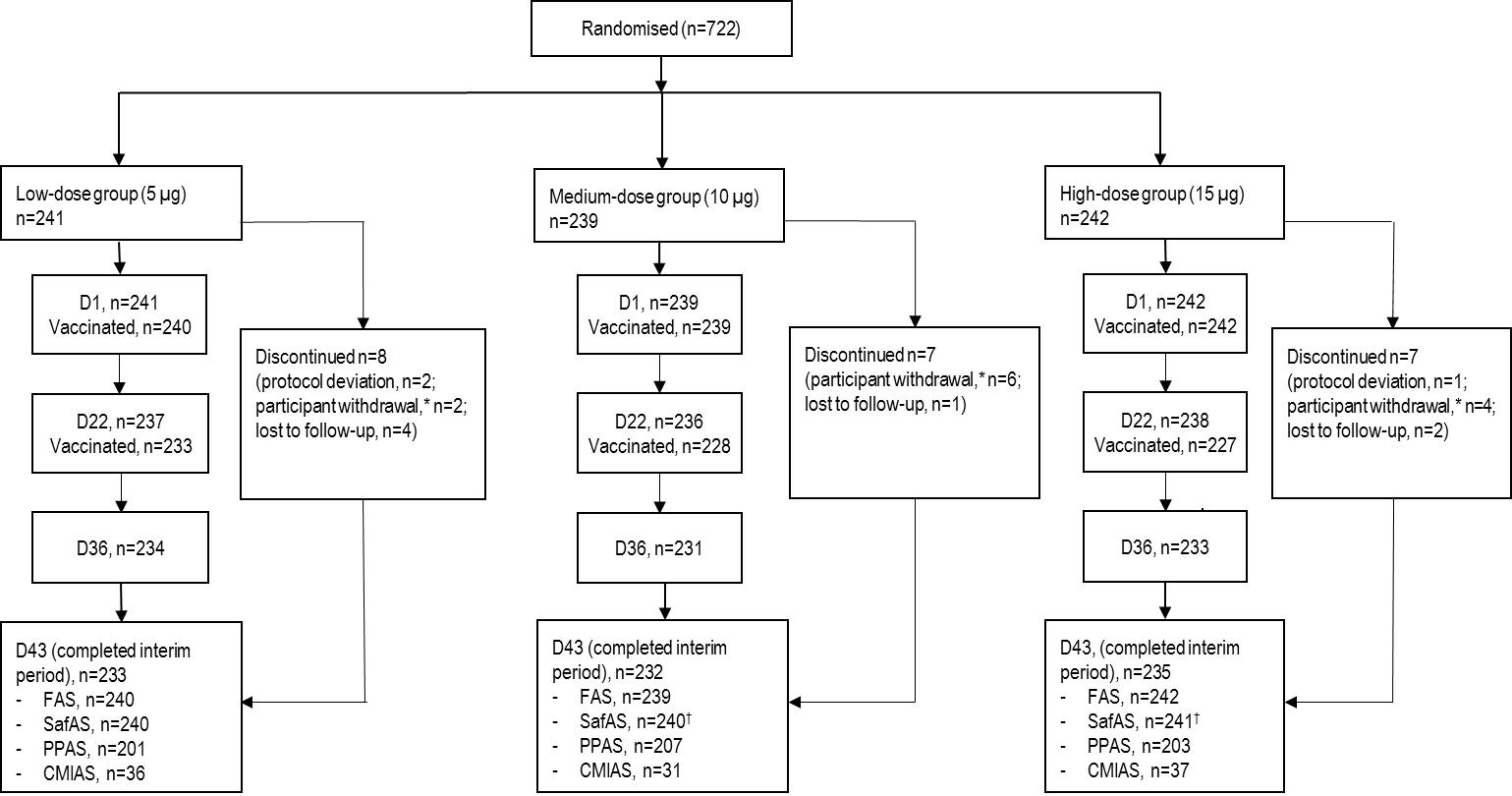

D = study day. CMIAS = cell mediated immunity analysis set. FAS = full analysis set. SafAS = safety analysis set. PPAS = per protocol analysis set.
*Not due to an adverse event; †One participant randomized to the high-dose group received medium antigen dose vaccine formulation on D0 and received a high-dose formulation as planned on D22.

### Figure S2. Solicited injection site and solicited systemic adverse reactions occurring over the first seven days after the second vaccination for younger adults (A; 18–59 years) and older adults (B; ≥60 years)

A. Low- (5 µg), medium- (10 µg), and high-dose (15 µg) groups - 18–59 years

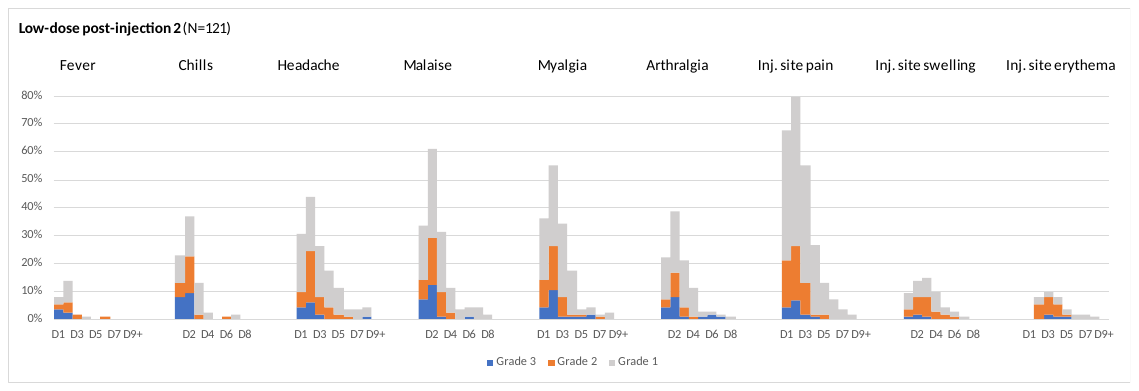

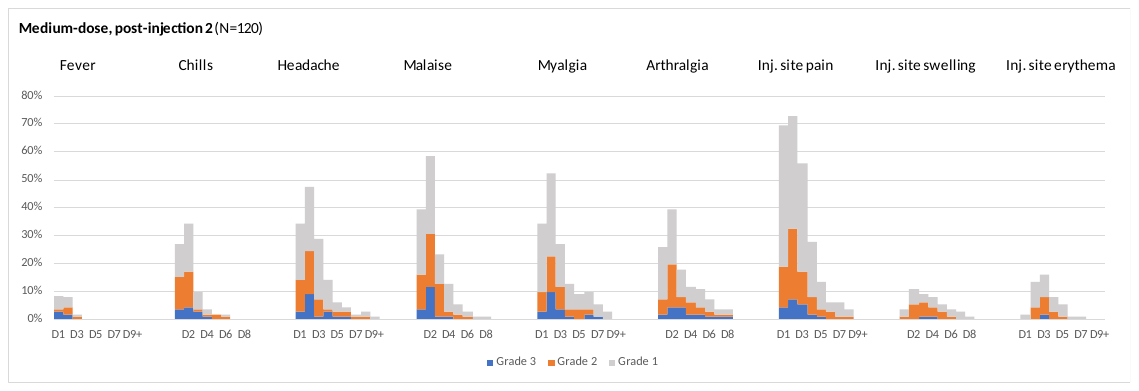

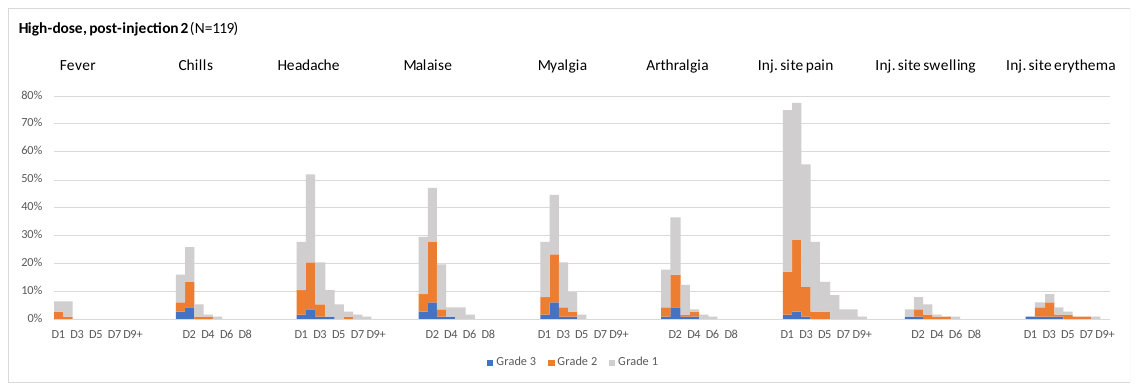

B. Low- (5 µg), medium- (10 µg), and high-dose (15 µg) groups - ≥60 years

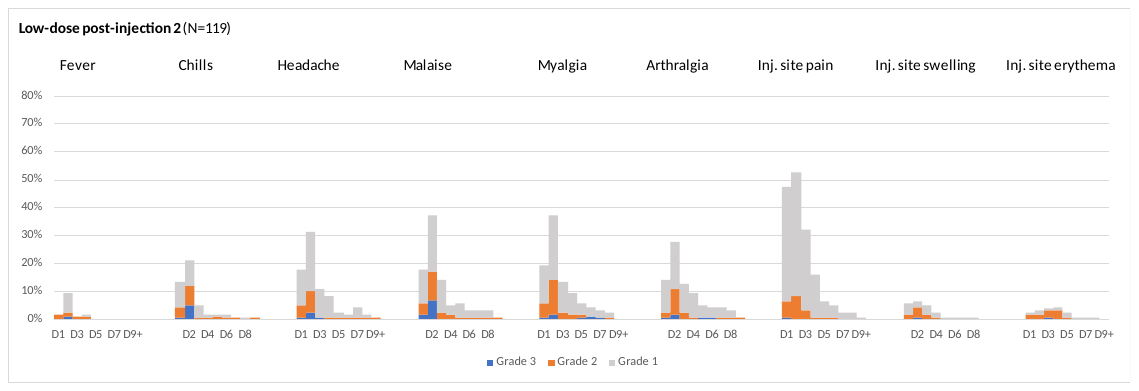

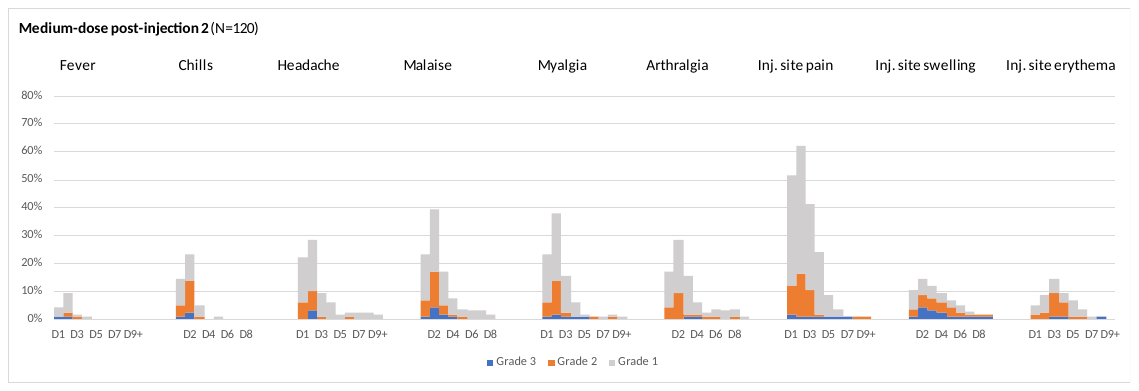

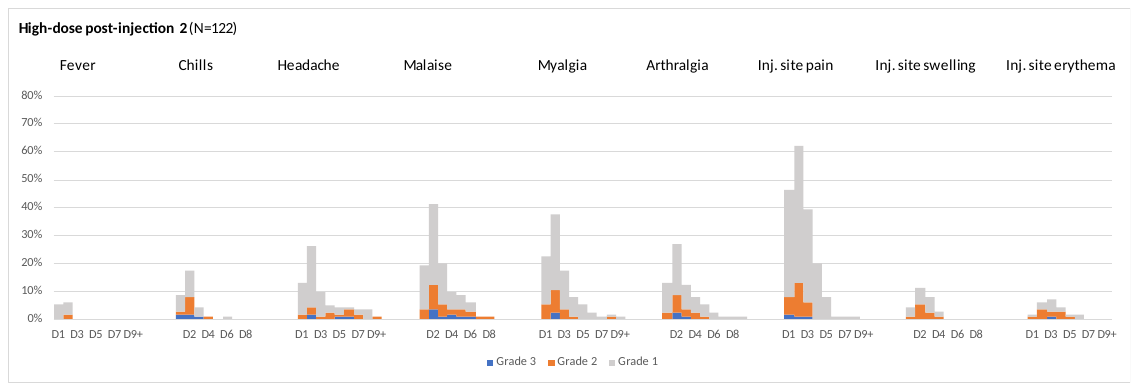

### Figure S3. Neutralising antibody response to the Beta variant, 14 days after the second injection of CoV2 preS dTM-AS03 (all dose groups combined), by SARS-CoV-2 naïve status Naïve = the subset of PPAS participants who were SAR-CoV-2-naïve at D1 and D22; N=521 Non-naïve = the subset of PPAS participants who were SAR-CoV-2-non-naïve at D1 and/or D22, N=77

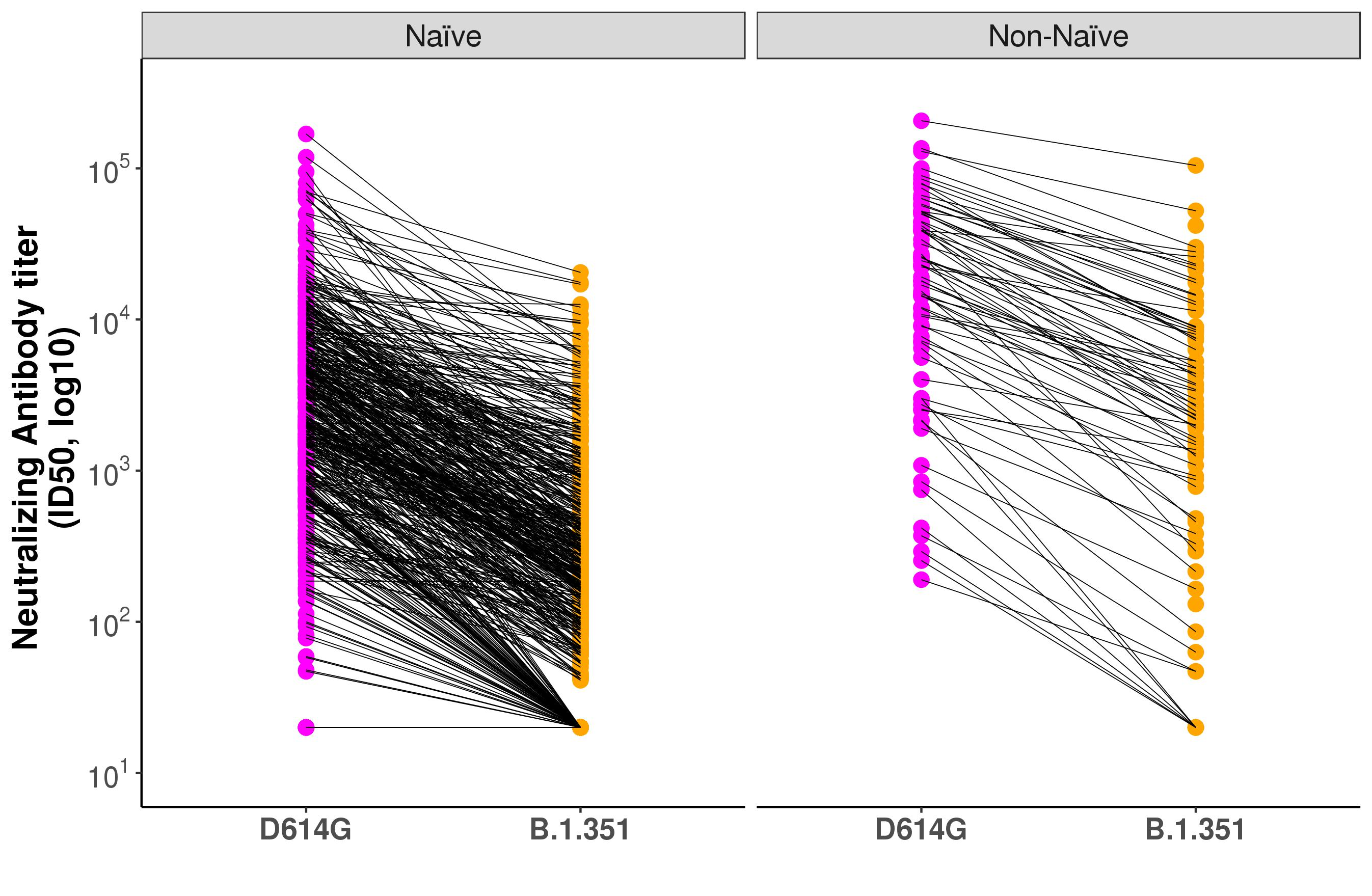

### Figure S4. CMI analyses – ratios of fold-rises for IFN-γ (A), IL-2 (B) and TNF-α (C) to IL-4, IL-5 and IL-13 at Day 22 and Day 36 (CMIAS)

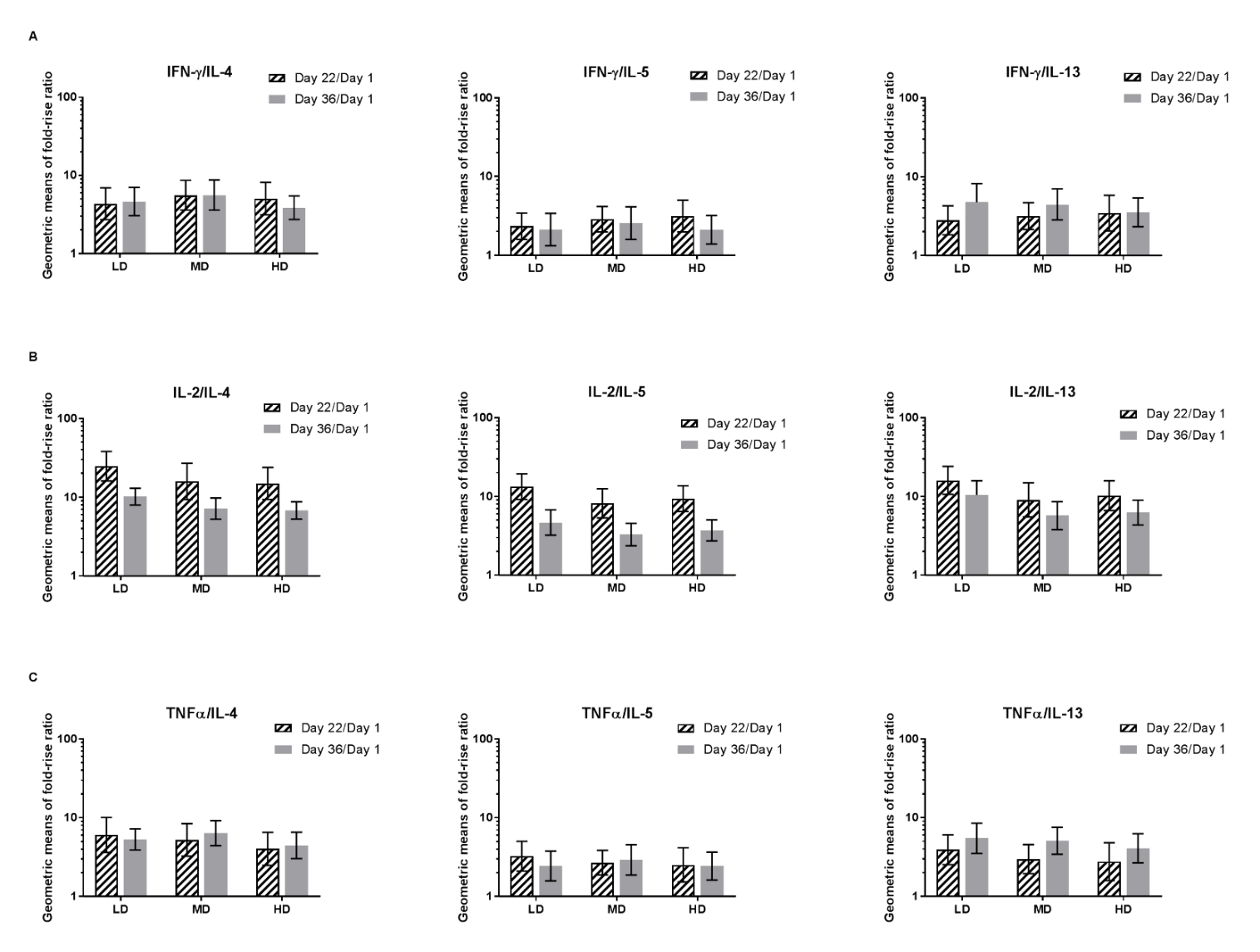

CMI = cell mediated immunity. CMIAS = cell mediated immunity analysis set. HD = high-dose (15 µg). IFN = interferon. IL = interleukin. LD = Low-dose (5 µg). MD = medium-dose (10 µg). TNF = tumour necrosis factor.
